# Epistatic contributions to human traits via transcription factor mechanisms

**DOI:** 10.1101/2025.09.28.25336826

**Authors:** Breeshey Roskams-Hieter, Olivier Labayle, Kelsey Tetley-Campbell, Mark J. van der Laan, Chris P. Ponting, Sjoerd V. Beentjes, Ava Khamseh

## Abstract

Epistasis causes an individual’s genetic background to modulate a DNA variant’s effect on trait [1–6]. Epistatic interactions among different loci in human complex traits are expected to be widespread but have not been found [7]. This could be due to small interaction effect sizes, the statistical complexity of estimating interactions that is higher than marginal variant effects, and a substantial multiple testing burden in a genome-wide scan [8–11]. Targeting interacting variants that contribute to the same biological pathway could lighten this burden. Here we combined Targeted Machine Learning [12, 13] with experimentally verified differential binding variants across 9 nuclear hormone receptors (NHR) to identify 535 two-point DNA variant-variant and 185 three-point variant-variant-sex NHR interactions among 768 traits in the UK Biobank (UKB) at a false discovery rate per trait of less than 0.05. Significance testing combined k allele-specific components into a Hotelling’s *T*^2^ test of Average Interaction Effect estimates at pairs/triples of loci (*k* ≤ 4 or *k* ≤ 8 for 2- or 3-point interactions, respectively). Nearly a third of 2-point interactions replicated, as they involved the same DNA-binding site and human trait but different trans-acting DNA variants. These epistatic mechanisms of altered transcription factor binding provide both plausible molecular mechanisms of action, and insight into sex-biased genetic risk, for diverse human traits and diseases.

## Main

A reduced statistical power of detecting DNA variant interactions does not necessarily imply that their true biological effect is smaller than that of single-variant effects. Epistatic interactions are fundamentally more difficult to statistically estimate than effect sizes of single variants even if, biologically, the former has a similar order of effect (Fig. 1A; Methods). When the true effect sizes of a 2-point Average Interaction Effect (AIE; Methods) and the 1-point Average Treatment Effect (ATE; Methods) [14, 15] are equal, for example, the power to detect the AIE is less than the power to detect the ATE at fixed sample size (Fig. 1A, middle). By contrast, when the true effect size of the AIE is half that of ATE, the power to detect AIE is substantially lower than the power to detect ATE (Fig. 1A, left). Even when the true effect size of the AIE is twice that of ATE, the power to detect AIE is at best equal to the power to detect ATE, but only when the population frequency of the heterozygous and rarest genotype of the interacting variant is 0.5. When the variant has a more typical genotype frequency, the power of detection drops quickly.

**Fig. 1:**
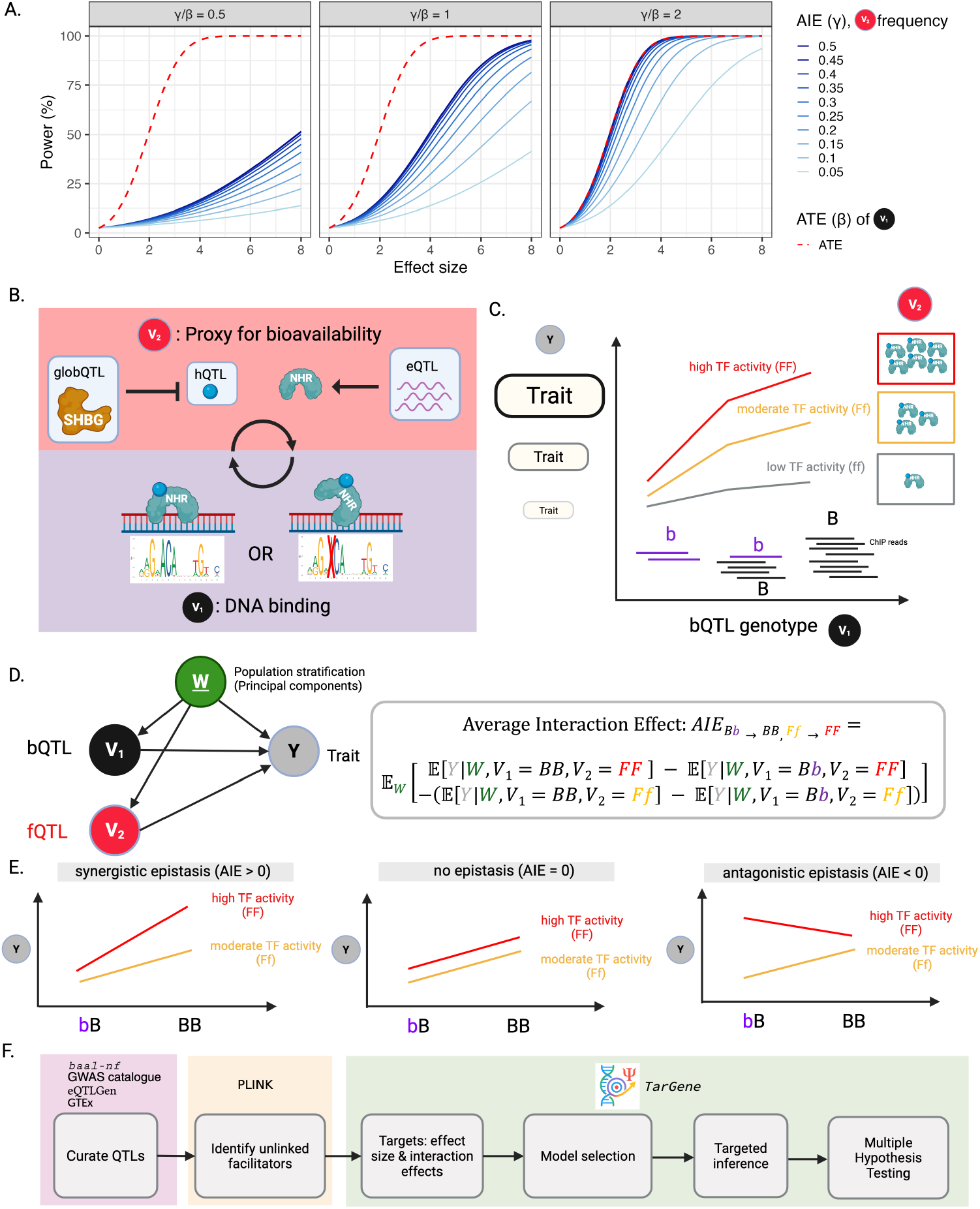
Proposed model of epistasis by altered transcription factor binding. **(A)** Statistical power of detecting average interaction effect (AIE; blue) of *V*_1_ and *V*_2_ on trait and average treatment effect (ATE; red) of DNA variant *V*_1_ on trait as a function of effect size in units of standard error. Different shades of blue represent different allelic frequencies for DNA variant *V*_2_. Power is displayed when the true magnitude of AIE (*γ*) is half as large as the true magnitude of ATE (*β*; left), equally large (middle), and twice as large (right). With a population frequency *f*_2_ = 0.5, detecting AIE with the same power as ATE requires either four times the sample size, or its effect to be twice as large. When *f*_2_ = 0.05, detecting AIE with the same power as ATE requires either ≈ 21 times the sample size, or its effect to be ≈ 4.59 as large. **(B)** The targeted epistatic mechanism by which altered binding at one locus can be modulated not only by a variant *V*_1_ that directly increases transcription factor binding affinity at this site (binding QTL; bQTL), but also by another, trans-acting variant *V*_2_ that facilitates modulation of TF activity (facilitator QTL; fQTL). We focus on a subset of TFs belonging to the nuclear hormone receptor (NHR) family depicted as blue proteins. We consider three types of fQTLs: expression QTL (eQTL), hormone QTL (hQTL) or globulin QTL (globQTL). For the remainder of the paper, we annotate plots with these fQTL labels. **(C)** We denote the biallelic genotypes of the fQTL *V*_2_ by ff, fF, and FF where F or f represent the “facilitator alleles” that increase or decrease, respectively, facilitation of TF activity. Here F increases abundance of our ligand-bound, activated NHR and its downstream consequence on trait is indicated graphically on the y-axis. Similarly, the biallelic genotypes of the bQTL *V*_1_ are bb, bB, and BB, where B represents the “binding allele” that increases transcription factor binding affinity at this site. **(D)** Left: Directed acyclic graph (DAG) displaying variables and their relations, where *V*_1_ denotes the DNA variant that modulates binding (bQTL), *V*_2_ denotes the DNA variant that modules TF activity via a facilitator (fQTL), and *Y* denotes trait. We account for genetic structure and population stratification by conditioning on principal components *W* computed across genotyped SNPs in UKB. Right: The model-independent formula for the AIE representing the epistatic interaction of bQTL *V*_1_ and fQTL *V*_2_ on trait *Y*, accounting for confounders *W* . Specifically, this AIE estimates the synergistic (or antagonistic) effect on trait of changing *V*_1_ from bB → BB and *V*_2_ from fF → FF simultaneously (Methods). **(E)** Illustrations of synergistic and antagonistic epistasis as a difference in conditional effects (relative slopes). In synergistic epistasis, the effect of the bQTL *V*_2_ on trait *Y* is greater at high TF activity [FF genotype] compared to moderate TF activity [fF genotype]. Conversely, antagonistic epistasis occurs when higher TF activity [FF genotype] results in a lesser effect of *V*_2_ on trait *Y* compared to moderate TF activity [fF genotype]. If there is no difference in the effect of bQTL *V*_1_ on trait *Y* between genotypes FF and fF, no epistasis is occurring. **(F)** Workflow for identifying epistatic interactions across complex traits in the UKB, including data curation, QC, and TarGene’s workflow: Statistical target definition, nuisance parameter estimation via model selection, targeted inference and multiple hypothesis testing correction.

The likelihood of detecting epistatic interactions can be enhanced by targeting specific interactions within particular molecular mechanisms, here involving 9 nuclear hormone receptors (NHRs; Table 1). These transcription factors (TFs) contribute to a wide diversity of physiological processes, and their gene variants have been associated with various human diseases [16]. NHR genetics provides an excellent paradigm for studying intermolecular epistatic interactions [17] due to (i) their two-step regulation of gene expression – typically involving an initial activation by a hormone ligand, followed by binding to DNA sites (“hormone-response elements”) near target genes [18–20], (ii) their involvement in diverse physiological processes [21], and (iii) their sexually dimorphic expression and function [22].

We chose to target a high-quality set of 119 variants (“binding quantitative trait loci”, bQTLs) that alter NHR binding affinity, and that are candidates for altering trait-causal mechanisms [23]. These bQTLs show significant allelic imbalance at a known heterozygous site in ChIP-seq reads (Fig. 1C) within a well-defined binding motif located in a ChIP-seq peak (Table 1, **Supplemental Table S2**). Additionally, we stringently required these alleles be concordant in their changes in binding affinity and in motif score [23]. Such ‘high quality’ bQTLs tend to be evolutionarily better conserved, have a higher number of known associations with traits, and a greater colocalization with molecular QTLs [23]. We tested for epistatic interactions with these bQTLs a set of trans-acting variants that modulate their NHR’s activity via bioavailability either of the NHR’s mRNA (expression QTLs; eQTLs) or of the hormone (hormone QTLs; hQTLs), including via sequestration of sex hormones by sex hormone binding globulin (SHBG; globulin QTLs, or globQTLs) (Fig. 1B, **Supplemental Table S1**). These biallelic trans-acting variants are facilitatory variants (fQTL) with alleles F and f, where F denotes the higher abundance of the trans-actor (and consequently, activity of the NHR). Similarly, bQTLs have alleles B and b, where B is associated with stronger DNA-binding affinity of the NHR (Fig. 1B,C). Facilitatory variants were not in strong linkage disequilibrium (LD; *R*^2^ *<* 0.8).

Methodologically, we sought to ensure estimates benefit from maximal statistical power and to minimise bias due to model misspecification. This is critical given our method’s application to a large-scale cohort (768 UKB traits [24]) and that effect sizes could be small. To this end, we applied TarGene, a statistical estimation technique and software founded on the semi-parametric estimation framework of Targeted Learning [12, 25, 26]. TarGene provides unbiased estimates of variant effects, 2-point interactions, and 3-point interactions with appropriate coverage of ground truth in large-scale realistic simulations modelled on UKB genotype-phenotype data [27, 28].

The Average Interaction Effect (AIE; Fig. 1D; Methods) defines an epistatic interaction between DNA variants *V*_1_ and *V*_2_ with respect to a trait *Y* whilst correcting for population structure as captured by principal components *W* . It can be interpreted as a difference between two conditional effects: The effect on trait of a change in bQTL at high TF activity [FF genotype] minus the effect on trait of a change in bQTL at lower TF activity [Ff genotype]. The interaction is synergistic when the AIE is positive: higher levels of TF activity [FF genotype] result in a greater effect of the bQTL on trait compared to lower levels of TF activity [fF genotype]. Contrarily, the interaction is antagonistic when the AIE is negative: higher levels of TF activity [FF genotype] result in a lesser effect of the bQTL on trait compared to lower levels of TF activity [fF genotype] (Fig. 1E, Methods). This is consistent with definitions of antagonistic and synergistic epistasis in the literature [5, 29]. We calculate the AIE as single-allelic transitions relating to increased levels of binding and facilitator activity, calculating 4 such pairs when genotypes pass the positivity criterion 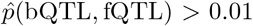 (Methods) [27, 28]. These *k ≤* 4 components are then tested at the bQTL-fQTL level using a Hotelling’s *T* ^2^ test [27, 28] which reduces the multiple testing burden relative to testing all allelic changes separately (Supplementary Material). Resulting p-values (termed *p*_*H*_ ) are adjusted using the Benjamini-Hochberg procedure and significant AIEs defined at a false discovery rate (FDR_*H*_ ) *<* 0.05 per trait per NHR (Methods).

### Replicated 2-point interactions for the progesterone receptor

Estimating 2-point interactions for each of the 9 NHRs, across 119 bQTLs and 147 fQTLs, yielded 535 significant bQTL-fQTL interactions for 768 UKB traits at an FDR_*H*_ *<* 0.05 (Methods, Fig. S5, **Supplemental Table S3**). Of these, 173 interactions (32.3%) involved an identical bQTL-trait pair, but different fQTLs. Such replicated interactions were discovered for 7 NHRs: PGR, AR, ESR1, NR2F1, NR2F2, HNF4A and HNF4G (Table 2).

As an example, we consider the 2-point significant interaction (*p*_*H*_ = 2.0 *×* 10^−4^, with FDR_*H*_ *<* 0.05) between a PGR eQTL (rs35802934 on chromosome chr 11q22.1) and a PGR bQTL (rs12626817 on chr 21q21.1) for ‘fracture head & neck’. In this case, Hotelling’s *T*^2^-statistic consists of *k* = 3 constituent AIE estimates (Fig. 2A, left); the fourth AIE (bB → BB; fF → FF) had insufficient frequency of the [BB,FF] genotype in the UKB population (Methods). Adding an additional PGR binding-increasing allele to [bB] for individuals with moderate expression of PGR [fF genotype] has a lower effect than the same addition in individuals with low expression of PGR [ff genotype], *i*.*e*., it produced an antagonistic interaction for the trait ‘fracture head & neck’ (blue, *p* = 8.28 × 10^−5^). In other words, the effect of the bQTL on ‘fracture head & neck’ is reduced for individuals with [fF] relative to those with [ff]. In contrast, adding these alleles to two other baseline genotypes ([bb,ff] or [bb,fF]) did not change this bQTL’s effect on associated disease risk for this trait (*p* = 0.52 and 0.84, respectively) (Fig. 2A). This is an example of idiosyncratic epistasis, when the effect of a mutation on trait depends on the genetic background and the type of interactions involved [30].

**Fig. 2:**
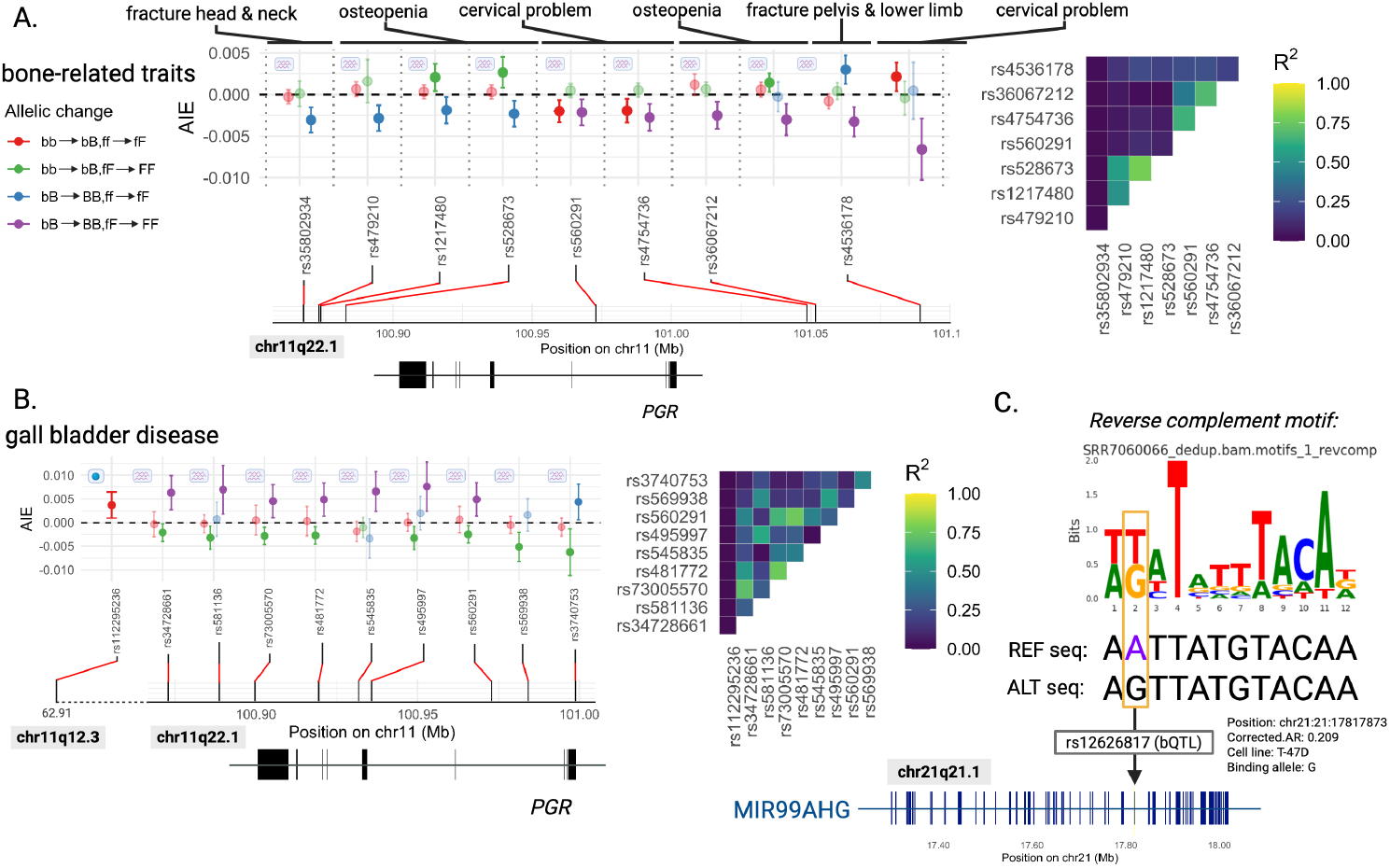
Two-point AIEs for the progesterone receptor (PGR) at rs12626817 are well-replicated across multiple traits. Figures **(A)** and **(B)** show allelic effects for significant interactions called after Benjamini-Hochberg correction on Hotelling’s *T* ^2^ test (*p*_*H*_ ) for PGR (FDR_*H*_ *<* 0.05). Y-axis shown represents the average interaction effect (AIE) size and associated 95% confidence intervals for individual genotype changes of the binding QTL and facilitator QTL. Non-significant individual effect sizes that cross the zero-line are shown at a higher transparency than those with *p <* 0.05. Lowercase b represents the non-binding allele and uppercase B represents the binding allele (ie. increases TF binding affinity). The facilitator allele is represented by f and F, where uppercase F represents an increase in the facilitator and f represents a decrease. In the context of eQTLs, for example, F indicates the allele that increases mRNA expression of the TF. **(A)** Interactions including the bQTL rs12626817 across 8 *PGR* eQTLs. None of these are in high LD with one another (*R*^2^ *>* 0.8), and most are in low LD (*R*^2^ *<* 0.25), as indicated by the heatmap of pairwise LD scores (*R*^2^). **(B)** For ‘gall bladder disease’, ten independent facilitator QTLs showed evidence for epistasis with the same bQTL, rs12626817, on the same trait. Each of these 10 interactions was significant after Benjamini-Hochberg correction on the Hotelling p-value at FDR_*H*_ *<* 0.05. Pairwise LD scores for each pair of facilitator QTLs are also shown in a heatmap. **(C)** rs12626817, the bQTL involved in these significant interactions, disrupts a *de novo* TF binding motif discovered by *baal-nf*. This SNP, located on chromosome 21 at position 17,817,873 [hg19] lies within an intronic region of the *MIR99AHG* gene. The allele that increases binding affinity at this site is the alternate (ALT) allele, G. The decrease in binding allele is the reference (REF) allele, A.

For the same bQTL and 3 other bone-related traits (‘osteopenia’, ‘cervical problem’ and ‘fracture pelvis and lower limb’), each of 7 *PGR* eQTLs showed significant 2-point interactions (Fig. 2A). These eQTLs are not in strong LD (*R*^2^ *<* 0.8) and for many, LD is weak (*R*^2^ *<* 0.4; Fig. 2A). Hence, these associations are largely statistically independent. They are also strikingly concordant. Adding a single PGR binding-increasing allele to [bb] for individuals with low expression of *PGR* [ff genotype] had no difference in effect for 7/10 of these bone diseases (red data points, 0.07 *< p <* 0.52). Interactions where single PGR binding-increasing and single *PGR*-expressing alleles were added to [bb,fF] (green), however, were synergistic for osteopenia for three *PGR* eQTLs (*p* = 4.89 × 10^−3^, 1.10 × 10^−2^, 1.36 × 10^−2^). In contrast, interactions where these same alleles were added to the [bB,ff] (blue) and [bB,fF] (purple) genotypes were largely antagonistic across ‘fracture head & neck’, ‘osteopenia’, ‘cervical problem’, and ‘fracture pelvis & lower limb’ and 8 *PGR* eQTLs (8.28 × 10^−5^ *< p <* 1.90 × 10^−2^) (Fig. 2A). Overall, adding [b,f] to alleles associated with higher PGR-binding and *PGR* expression tended to reveal antagonistic interactions (blue and purple), whereas their addition to lower PGR-binding and *PGR* expression alleles tended to yield more synergistic interactions (green).

Thirty-three other traits were significantly associated with epistatic interactions with this bQTL and one or more of these progesterone hQTLs or *PGR* eQTLs (**Supplementary Table S3**). ‘Gall bladder disease’ yielded significant epistatic interactions between this PGR bQTL and each of 9 *PGR* eQTLs or an hQTL (FDR_*H*_ *<* 0.05; Fig. 2B). Compared to the bone disorders in Fig. 2A, ‘Gall bladder disease’ showed an opposite direction of effect, with more synergistic interactions at higher binding and *PGR* expression alleles. These interactions were, again, remarkably concordant across trans-actors: where a single PGR binding-increasing and single *PGR* expression-increasing alleles were added to the [bb,fF] genotype (green) these were largely antagonistic across 8/10 *PGR* eQTLs (9.47 × 10^−4^ *< p <* 2.59 × 10^−2^), whereas adding such alleles to the [bB,fF] genotype (purple) yielded synergistic interactions across 7/10 *PGR* eQTLs (5.50 × 10^−4^ *< p <* 1.15 × 10^−2^) (Fig. 2B). Adding single PGR binding-increasing and single *PGR* expression-increasing alleles to [bb,ff] (red) did not show significant evidence of epistasis (0.08 *< p <* 0.98). However, adding a single progesterone-increasing allele to [bb,ff] (red) produced a synergistic interaction on this trait (*p* = 8.85 *×* 10^−3^) (Fig. 2B, left).

These epistatic interactions all involved the same PGR bQTL (rs12626817), whose higher-binding allele (G) strengthens the *de novo* PGR binding site motif discovered by *baal-nf* (Fig. 2C); it is also the derived allele, as other apes carry the lower-binding A-allele. This bQTL lies within an intron of the miR99a/Let7c/miR125b2 host gene, *MIR99AHG*. These 3 microRNAs are members of the evolutionarily ancient miR-10, let-7 and lin-4 families, respectively, which regulate diverse functions including osteogenesis, neuronal maturation, haematopoiesis, megakaryopoiesis, and cell proliferation and differentiation [31–35].

Next, we found 4 significant 3-point interactions involving PGR (**Supplemental Table S4**), 3 of which were synergistic. These involved both binary and continuous traits, and different transactor mechanisms (bQTL, hQTL and eQTL).

### Epistasis yields plausible biological mechanisms

The 2-point interactions for AR, NR3C1, VDR and ESR1 in Fig. 3B exemplify the diversity of TF mechanisms across traits, directions of effect, and candidate downstream targets. Replicated interactions for AR and bone-related disorders involved multiple fQTLs and the same bQTL. Adding single AR-binding and single testosterone-increasing alleles to the [bb,ff] genotype produced synergistic interactions (red; *p* = 7.03 × 10^−4^ and *p* = 4.84 × 10^−4^), whereas adding these same alleles to the [bb,fF] genotype produced antagonistic interactions (green; *p* = 4.67 × 10^−4^ and *p* = 8.02 × 10^−5^) (Fig. 3A). This bQTL’s T-allele (rs2553234) increases AR-binding affinity while strengthening the JASPAR AR motif MA0007.2 (Fig. 3B). As this bQTL is also a *cis*-eQTL for the *ANXA2* gene, the increase in AR-binding is also associated with increased *ANXA2* expression. Due to *ANXA2* ‘s known role in bone mineralization [36], our findings suggest that it contributes to these bone disorder traits epistatically through hormone-modulated altered AR binding at this locus.

**Fig. 3:**
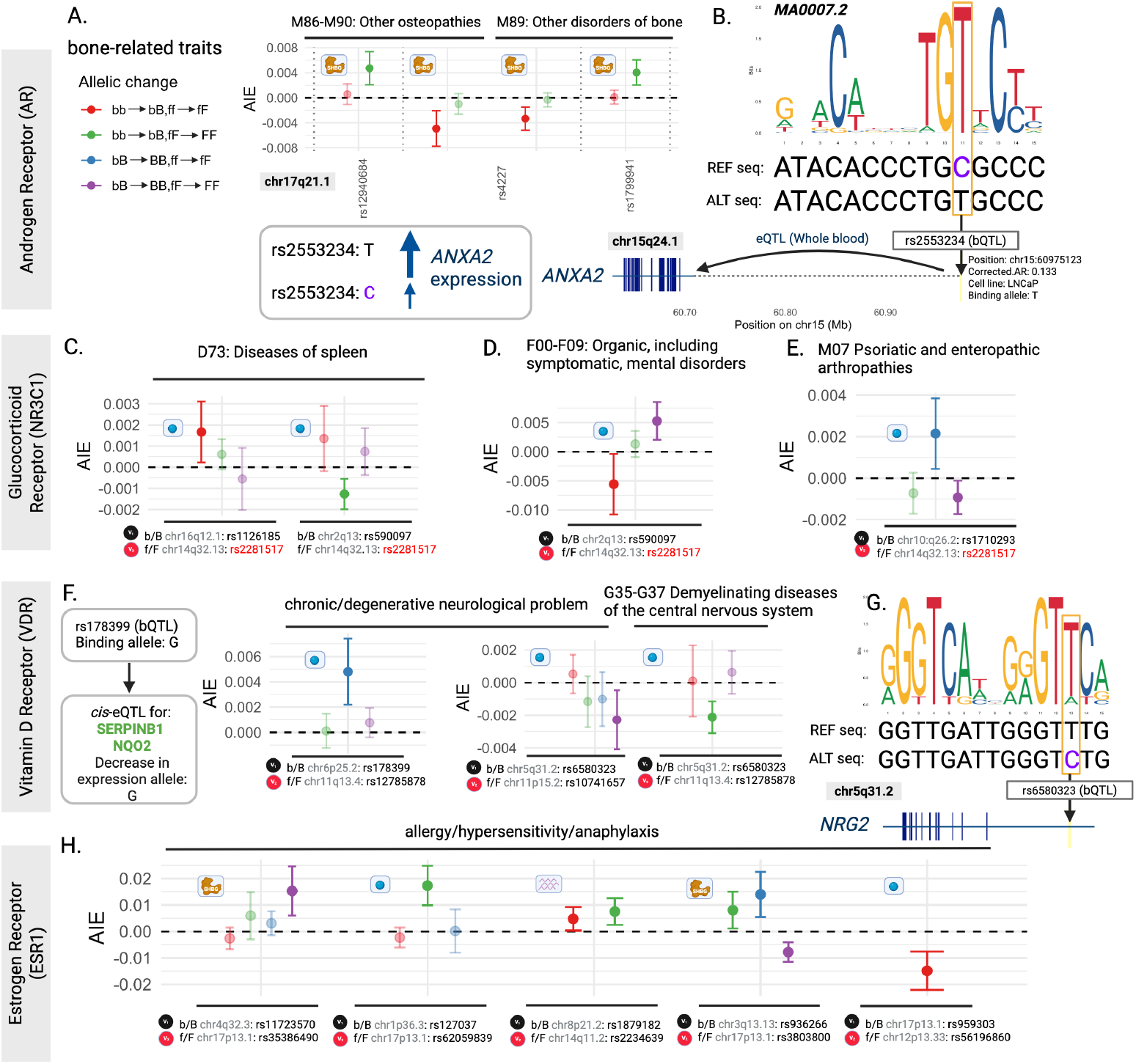
Epistatic mechanisms of altered TF binding provide plausible biological mechanisms of action. **(A)** Replicated interaction for the Androgen Receptor across two different bone disorder traits: (i) ‘Other osteopathies’ and (ii) ‘Other disorders of bone’. Three transactors (on the x-axis) interacted with the same bQTL, rs2553234, on both traits. For this bQTL, b represents the non-binding allele [C] and B represents the binding allele [T]. All three facilitators included in this interaction are protein QTLs for SHBG. Across these three SNPs, the f allele indicates a higher amount of SHBG protein, and therefore a lower amount of testosterone available to bind AR. Consequently, the F allele indicates lower amount of SHBG, which we use as a proxy for higher amounts of testosterone that can bind AR. Variation in AIE values could reflect differences in effect sizes across facilitators (**Supplementary Table S1**). **(B)** JASPAR motif for AR, MA0007.2, with the SNP position for rs2553234 (located on chr15q24.1) highlighted in a yellow box, showing that the increase in binding allele, T (ALT allele), increases the strength of this motif, and the decrease in binding allele, C (REF allele), disrupts the motif. This bQTL is also an eQTL in whole blood for *ANXA2* [37], where an increase in binding at this site is also associated with increased *ANXA2* mRNA expression. **(C-E)** Three examples of significant epistatic interactions found for the glucocorticoid receptor (NR3C1). All facilitators in these plots are for rs2281517, where f indicates the allele associated with lower cortisol levels and F indicates the allele associated with higher cortisol levels. For each interaction the bQTL is listed below the plot, along with chromosome and position, indicating that these interactions are in trans. Here we report interactions across **(C)** ‘Diseases of spleen’, **(D)** ‘Organic, including symptomatic, mental disorders’ and **(E)** ‘Psoriatic and enteropathic arthropathies’. Panel **(F)** includes significant 2-point interactions for the Vitamin D Receptor (VDR) across 2 bQTLs, rs178399 for the ‘trait chronic/degenerative neurological problem’, and rs6580323 for 2 traits, ‘chronic/degenerative neurological problem’ and ‘Demyelinating diseases of the central nervous system’. All fQTLs across these interaction effects represent hormone QTLs for 25-hydroxyvitamin D, the primary form of vitamin D circulating in the blood. rs178399 is an eQTL (Whole blood) for *SERPINB1* and *NQO2* [37]. **(G)** rs6580323 disrupts the JASPAR motif for VDR, MA0074.1, and lies within the intronic region of *NRG2*, a gene that promotes neuronal survival and neurite extension [38]. **(H)** Two-point AIEs discovered for ESR1 across 5 bQTLs and 5 fQTLs for the same trait, ‘allergy/hypersensitivity/anaphylaxis’. One of these bQTLs, rs936266, is also an eQTL for *CD47*, which is involved in Th2-mediated allergic inflammation [39]

For NR3C1, diseases of spleen (ICD-10 code D73) were involved in 2 epistatic interactions with the same fQTL (for cortisol levels) and 2 different bQTLs (Fig. 3C). For this trait, AIE direction of effect varied with increasing binding or hormone levels. For bQTL rs1126185, addition of single NR3C1 binding and single cortisol-increasing alleles to [bb,ff] was synergistic for this trait (red; *p* = 2.35 × 10^−2^). Conversely, addition of single NR3C1 binding and single cortisol-increasing alleles to [bb,fF] for bQTL rs590097 was antagonistic. This indicates that the bQTL’s effect on trait was significantly reduced for individuals with genotype [FF] compared to those with genotype [fF] (green, *p* = 5.78 × 10^−4^). The 2 bQTLs highlighted in Fig. 3C are eQTLs for *WWOX* non-coding and/or *ACOXL* protein-coding transcripts in the spleen and brain [40], the 2 affected tissues. Cortisol is a biomarker of mental disorder severity [41], and is used to treat psoriatic and enteropathic arthropathies [42], 2 traits for which NR3C1 bQTL interactions were significant (Fig. 3D,E).

For VDR, ‘chronic/degenerative neurological problem’ was involved in significant 2-point interactions for 2 bQTLs, with opposite directions of effect (Fig. 3F). All fQTLs investigated in these interactions represented an increase in 25-hydroxyvitamin D (250HD), the primary form of vitamin D circulating in the blood. One of these VDR bQTLs, rs6580323, was also involved in a significant 2-point interaction with ‘G35-37 Demyelinating diseases of the central nervous system’. Adding a single VDR binding allele to [bb] or [bB] for individuals with high levels of circulating vitamin D [genotype FF] had a lesser effect on these neurodegenerative diseases compared to the effect of these bQTL changes in individuals with moderate levels of circulating vitamin D [genotype fF] (*p* = 2.18 × 10^−5^ and *p* = 1.36 × 10^−2^ for [bb] (green) and [bB] (purple), respectively; Fig. 3F). This bQTL disrupts the JASPAR motif for the RXRA::VDR heterodimer, MA0074.1, and lies within an intron of *NRG2* (Fig. 3G), a gene that promotes oligodendrocyte survival and function and is important in myelin maintenance [43].

For ESR1, a single trait ‘allergy/hypersensitivity/anaphylaxis’ showed evidence of epistasis across 5 bQTLs and 5 fQTLs (Fig. 3H). These interactions occur across all three transactor mechanisms (*ESR1* expression, estradiol abundance and SHBG abundance) and showed variable directions of effect and contributions of component genotype change. Estrogen signalling impacts allergies and hypersensitivities in humans and model organisms [44–47].

Both HNF4A and HNF4G had significant interactions involving the same two bQTLs: rs514795 and rs6470263 (**Supplemental Table S3**, Fig. S5A). This is expected because these 2 paralogues share binding sites [48, 49], can form heterodimers [50], and have partially overlapping functions [51]. However, HNF4A and HNF4G did not share significant epistatic interactions. For the rs514795 bQTL, our study associated HNF4A with eye traits (‘H25 senile cataract’, ‘H25-H28 Disorders of lens’, and ‘eye trauma’), for example, consistent with *Hnf4a* knockout mice (*Hnf4a*^*tm*1*b*(*EUCOMM*)*Hmgu*^) exhibiting a cataract phenotype, and HNF4A modulating the epithelial-mesenchymal transition of lens epithelial cells [52]. We also associated *HNF4A* with diabetes consistent with its mutation in one form of diabetes [53]. In contrast, our study associated the rs514795 bQTL and *HNF4G* with ‘M07 Psoriatic and enteropathic arthropathies’, a broad code that includes enteropathic arthritis often associated with inflammatory bowel disease, whereas we associated rs514795 and *HNF4A* with the overlapping code ‘K50 Crohn’s disease [regional enteritis]’. This disease overlap is consistent with the similar, but non-identical, expression pattern of *HNF4A* and *HNF4G* in the intestine [54, 55]. These findings demonstrate this approach’s ability to discover interpretable and biologically meaningful epistatic interactions across diverse human traits.

### Replicated sex-differential interactions on diverse traits

A bQTL-fQTL interaction on trait could be additionally modulated by sex (Fig. S6). For 5 NHRs with a well-defined hormone ligand (AR, PGR, ESR1, VDR and NR3C1), and across 114 fQTLs and 108 bQTLs, we identified 185 significant 3-point interactions involving bQTL, fQTL and genetic sex (FDR_*H*_ *<* 0.05) (**Supplemental Table S5**, Fig. S7). Twenty five (13.5%) of these 3-point interactions were replicated, and involved the same PGR, AR or ESR1 bQTL-trait pair across multiple fQTLs. (Table 3). A 3-point interaction is positive when the bQTL-fQTL interaction on trait is larger among males than females (see Eq. 3, Methods). This is interpreted as the interaction effect being (i) more synergistic in males than in females, or (ii) synergistic in males and antagonistic in females, or (iii) less antagonistic in males than in females.

For each significant trait-bQTL-fQTL 3-point interaction with sex, we also estimated 2-point (bQTL-fQTL) interactions on the trait-of-interest for males or females, highlighting those showing significant differences between sexes (FDR_*H*_ *<* 0.05). This provided directionality of each epistatic interaction per trait, separated by sex.

‘Viral agents as the cause of diseases classified to other chapters’ (ICD-10: B97) is an aggregate trait describing diseases of viral origin. For each of 2 fQTLs in low LD (*R*^2^ = 0.21), the addition of a single PGR binding-increasing allele at rs12626817 to [bb] had a greater effect on this disease trait for females with high expression of *PGR* [FF genotype] compared to females with moderate expression of *PGR* [Ff genotype]. This interaction was synergistic in females (green circles), but either antagonistic or absent in males (green triangles) (Fig. 4A). The relevant bQTL lies within the miR-99a, Let-7c and miR-125b-2 host gene (*MIR99AHG* ) whose miRNAs, and/or their related family members, regulate viral replication [60–65] and have known sex-biased functions [66] including due to hormone regulation [67] (Fig. 4C).

**Fig. 4:**
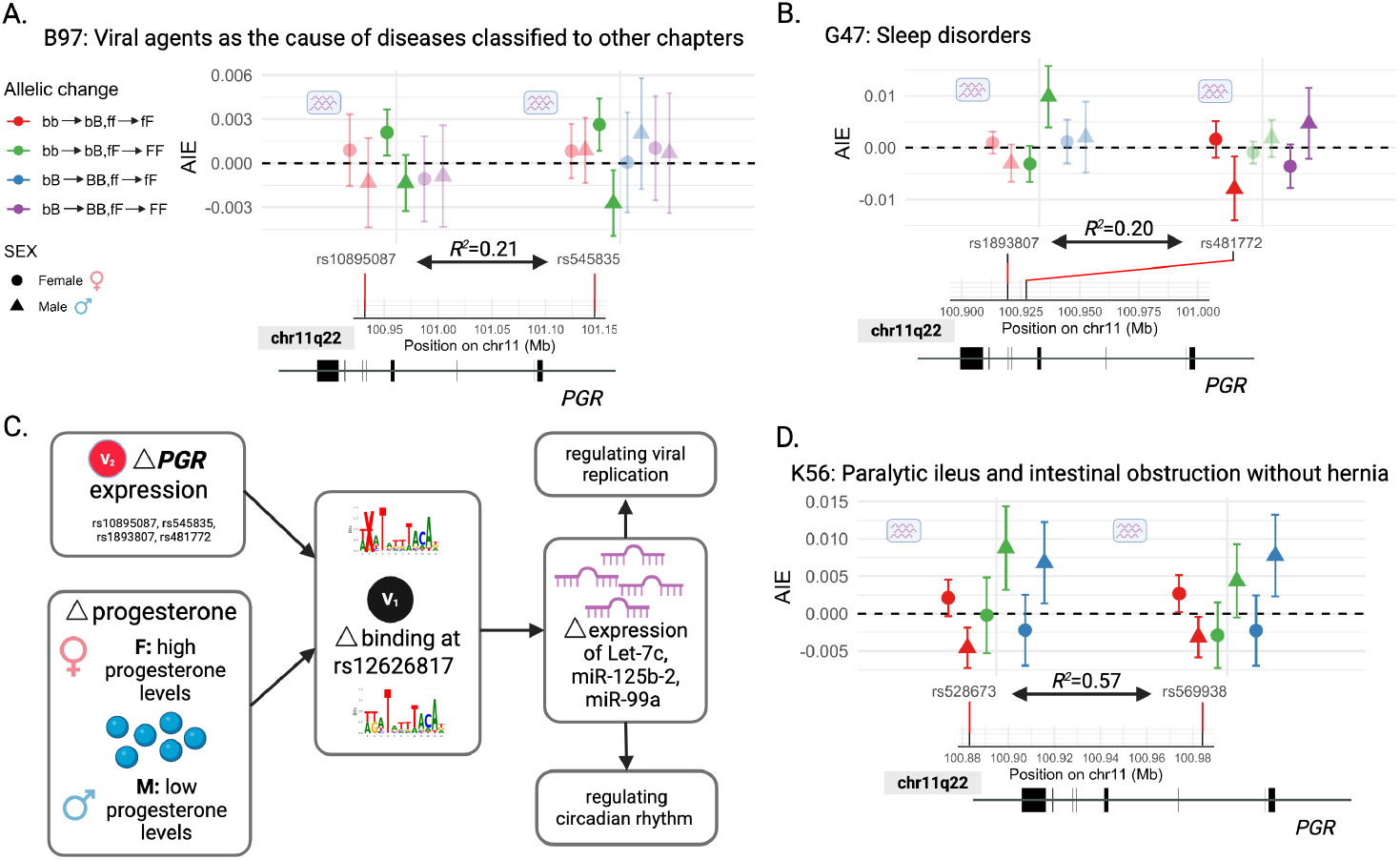
Replicated sex-biased 3-point interactions for PGR at rs12626817 involving diverse traits. Two significant (FDR_*H*_ *<* 0.05) epistatic interactions at rs12626817 for 2 facilitator QTLs with the same direction of effect are shown for: **(A)** ‘Viral agents as the cause of diseases classified to other chapters’, **(B)** ‘Sleep disorders’, and **(D)** ‘Paralytic ileus and intestinal obstruction without hernia’ (FDR threshold on Hotelling’s *T*^2^ statistic p-value, FDR_*H*_ *<* 0.05). Effect sizes for the individual bQTL/fQTL allelic changes and their 95% CIs are shown, for interactions involving males (triangles) or females (circles); interactions that are not significantly different between males and females are displayed at higher transparency. Both pairs of fQTLs reported across (A) and (B) are in low LD: *R*^2^ = 0.21 and *R*^2^ = 0.20, respectively. All fQTLs here are *PGR* eQTLs. The rs12626817 PGR bQTL lies within an intron of *MIR99AHG*, the host gene for 3 intronic miRNAs: miR-99a, Let-7c and miR-125b-2 (shown in Fig. 2C). **(C)** We propose that altered PGR binding at rs12626817 affects expression of miR-let-7c, miR-125b-2 and/or miR-99a. These miRNAs regulate viral replication [56–58], and circadian rhythm in Drosophila [59].

For ‘Sleep disorders’ (ICD-10: G47), the 2-point interaction (bb → bB; ff → fF; bQTL, rs12626817; fQTL, rs481772; red) was antagonistic (AIE *<* 0) for males (triangles) but not females (circles; Fig. 3B). Conversely, a different 2-point interaction (bb → bB; fF → FF; bQTL, rs12626817; fQTL, rs1893807; green) was synergistic (AIE *>* 0) for males but not females (Fig. 3B). One of the miRNA transcripts of *MIR99AHG*, Let-7, regulates circadian rhythm in Drosophila [59].

A similar pair of sex-biased 3-point interactions was discovered for the ‘Paralytic ileus and intestinal obstruction without hernia’ (ICD-10: K56) trait, involving 2 fQTLs in moderate LD (*R*^2^ = 0.57; Fig. 4D). Here, sex bias is evident when single PGR binding-increasing and *PGR* expression-increasing alleles are added to [bb,ff], [bb,fF] or [bB, ff]. The genotype [BB,FF] did not meet the positivity criterion and therefore is not shown. In males, adding these alleles to [bb,ff] was antagonistic (red triangles) for this trait, but synergistic when added to [bb,fF] or [bB,ff] (green or blue triangles, respectively) (Fig. 3D). These epistatic effects were not seen in females.

## Discussion

Evidence for epistasis for complex traits in human populations has been scant [7]. This study identified 535 two-point (bQTL-fQTL) interactions, 185 three-point (bQTL-fQTL-sex) interactions, and 17 three-point (bQTL-fQTL-fQTL) interactions involving NHRs at an FDR_*H*_ *<* 0.05 per NHR per trait (**Supplemental Tables S3 and S5**). Rather than parameterizing epistasis using variance components [7], we estimate 2- and 3-point interactions of genomic variants on polygenic traits in a targeted approach that maximises both statistical power and control over false discoveries [27, 28]. Results are population averages and thus reflect statistical rather than physiological epistasis [68]. Nevertheless, single individuals’ genotyped cell lines were used to define the bQTLs we tested for interactions [23, 69].

Twelve human DNA variants are highlighted in Figures 2-4 that strengthen or weaken NHR-binding affinity. Despite most new mutations being deleterious [70], 6 of these 12 variants gained binding affinity and did so recently, over hominin evolution (**Supplementary Table S6**). At one AR gain-of-binding site (rs2553234) the ancestral allele (A; present in chimpanzee, marmoset and dog) is absent from the modern human population; both human alleles (T or C) are, instead, derived. Vindija and Altai Neanderthal genomes carry the stronger (T) allele, and not the weaker (C) AR-binding allele which is the more frequent allele across modern human populations (gnomAD v4.1) (Fig. 3B). Relative to the C-allele, the lower frequency T-allele is protective of bone disorders in some genetic backgrounds (rs4227: interactions in red, Fig. 3A) but contributes risk in others (rs12940684, rs1799941: interactions in green, Fig. 3A). This reflects that both positive and negative interactions can occur between the same pair of variants, depending on whether these variants are heterozygous or homozygous [30, 71, 72].

Strengths of this study include (i) the large number of human traits investigated, (ii) the investigation of 2- and 3-point interactions, (iii) the ease of interpreting downstream consequences on genes, cells and tissues when a bQTL is also a known eQTL for neighbouring genes, and (iv) investigation and discovery of epistatic interactions in *trans*. Evidence from epistatic interactions should be at least as valuable as single human variant associations (*i*.*e*., 1-point genetic interactions) for drug discovery [73]. Our study investigated only a limited number of TFs (9 NHRs), and a low number of their bQTLs so as to improve statistical power of detecting these interaction effects.

As for any genetic association, confidence in these interaction results would increase if they were successfully replicated in an independent data set. The availability of TarGene [27, 28] facilitates future replication studies. Replication is expected to have greatest success in similar genetic ancestries. This is because variants of interest could be involved in higher order interactions with other variants whose allele frequencies vary across different ancestries. Nevertheless, when findings replicate then either other relevant variants will not be involved, or they will have similar allelic frequencies.

This study’s large set of predictions is consistent with widespread epistatic interactions in humans, which has long been suspected [74] but not demonstrated. How epistatic interactions can be interpreted is challenging [75]. To this end, we designed our study specifically to ease interpretation by linking TF-mediated, and hormone-modulated, molecular mechanisms to human traits and diseases via the TF’s bQTLs. For each interaction, downstream gene targets for involved bQTLs are identifiable either by (i) proximity of nearby genes or (ii) a known association between the bQTL and changes in gene expression, as identified in independent eQTL studies. The 3-point genetic effects could help discover sex-differential drugs and their dosage optimization. Future research should apply this study’s methods to all human TFs and traits in UK Biobank, as well as new cohorts, helped by the availability of human bQTLs [23] and the user-friendly software TarGene [27], to link diverse molecular mechanisms to human traits and disease susceptibility.

## Methods

### Model-independent definition of ATE and AIE

The genetic effect of variant *V*_1_ on trait *Y* is given by the Average Treatment Effect (ATE). The ATE is defined model-independently as

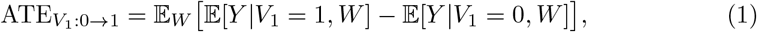

where *W* represents population stratification as a confounder (encoded by principal components), and labels 0 and 1 represent two genotypes, *e*.*g*., AA and Aa, or Aa and aa. The epistatic 2-point interaction between variants *V*_1_ and *V*_2_ on trait *Y* is given by the Average Interaction Effect (AIE). The AIE is defined model-independently as

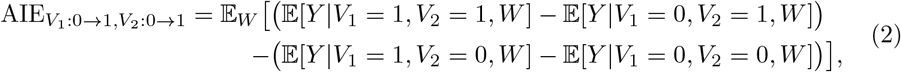

and displayed in Fig. 1D [76]. This definition has a natural interpretation as an extension of ATE: “Having adjusted for confounders, how does the effect of one variant *V*_1_ on trait *Y* change depending on the status of a second variant *V*_2_?”. This constitutes a difference between two conditional effects, namely the ATE of *V*_1_ on *Y* at *V*_2_ = 1 fixed (top line) minus the ATE of *V*_1_ on *Y* at *V*_2_ = 0 fixed (bottom line). The above model-independent definitions are entirely general, without imposing any restriction on the true functional form of dependence among the trait, variants, and population stratification. In the special scenario where the ground truth relationship is linear in each variable, *i*.*e*., *Y* = *α*_0_ + *α*_1_*V*_1_ + *α*_2_*V*_2_ + *γV*_1_*V*_2_ + δ*W*, then the model-independent definitions of ATE (Eq. 1) and AIE (Eq. 2) can be expressed in terms of the model’s coefficients. Specifically, the marginal effect of *V*_1_ on trait, also often referred to as the ‘*β*-coefficient’ (see Fig. 1A) equals 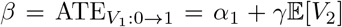. It involves a contribution from both the pure effect *α*_1_ of *V*_1_ on *Y* as well as the pair-wise interaction term *γ* and the genotype frequency 𝔼[*V*_2_] of *V*_2_. In this model, the interaction effect equals 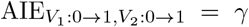, as naively expected. When the ground truth relationship between *Y* and (*V*_1_, *V*_2_, *W* ) is not linear in each variable, the expression of the ATE and AIE in terms of the model parameters generally differs from these equations. In the Supplementary Materials, we analytically demonstrate the lower statistical power of detecting AIE relative to the marginal ATE, at fixed total sample size *N* . We derive a closed-form expression for the statistical power in each case as a function of the sizes of these effects and genotype frequencies (see Fig. 1A).

In the context of NHRs and bQTL-fQTL interactions specifically, AIE constitutes the effect on trait of a change in bQTL at high fQTL level minus the effect on trait of a change in bQTL at low fQTL level (Fig. 1E). If the 2-point interaction is positive [AIE *>* 0], then high TF activity levels, denoted by the genotype FF, result in a greater effect of the bQTL change in trait, compared to lower TF activity levels, denoted by the genotype Ff, for example. If negative [AIE *<* 0], then high TF activity levels [FF genotype] reduce the effect of the bQTL change on trait, compared to lower TF activity levels [Ff genotype]. Since interactions are symmetric, this interpretation can be flipped around: The 2-point interaction is also the effect on trait of a change in fQTL at high binding level [represented by bQTL=BB] minus the effect on trait of a change in fQTL at low binding level [represented by bQTL=Bb].

Alternatively, bQTL-fQTL interactions can be interpreted as the difference between the effect on trait of changing both bQTL and fQTL [ATE(*V*_1_, *V*_2_)] relative to the sum of the effects on trait by a change in either bQTL or fQTL individually [ATE(*V*_1_) + ATE(*V*_2_)] (Fig. S2). If the 2-point interaction is positive [AIE *>* 0], the joint change of bQTL and fQTL is said to be “synergistic”, as it leads to a larger effect on trait than if both the fQTL and bQTL are changed individually. If the 2-point interaction is negative [AIE *<* 0, the joint change of bQTL and fQTL is said to be “antagonistic”, as it leads to a lower effect on trait than if both fQTL and bQTL are changed individually.

Two-point interactions have previously been studied in the experimental design and causal inference literature [29, 77–80]. This definition has been further generalised to higher-order (beyond pair-wise) interactions in our earlier work [27, 28, 76]. In this work, we examine 3-point interactions amongst variants *V*_1_, *V*_2_ and sex *S* on trait consisting of eight terms. It is defined model-independently as:

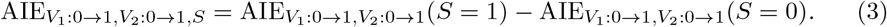

Here 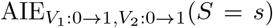 denotes the 2-point interaction in the subpopulation of individuals with sex *S* = *s, i*.*e*., the subpopulation of males (*S* = 1) or females (*S* = 0), namely

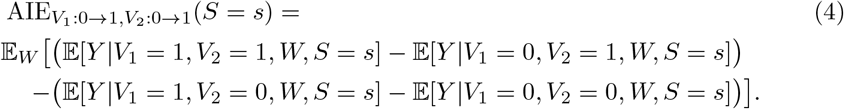

This 3-point AIE of variants *V*_1_, *V*_2_ and sex *S* (or a third variant *V*_3_) has the following interpretation: “Having adjusted for confounders, how does the interaction between *V*_1_ and *V*_2_ on trait *Y* change depending on sex (or the status of a third variant *V*_3_)?”. Similar interpretations exist for *n*-point interactions [76]. For details of the estimation pipeline, see Methods “Epistasis estimation with TarGene”.

### QTL curation and quality control

Across all analyses, our population-of-interest was the self-reported “White British” population in UKB (459,105 individuals). The set of 768 human traits investigated across this population matched those reported in GeneAtlas [24].

#### Candidate bQTL sets per NHR

Candidate binding quantitative trait loci (bQTLs) were identified using *baal-nf* [23]. Briefly, to be called a bQTL a DNA variant needed to meet three criteria: (i) it was a heterozygous SNP tested in genotyped human cell lines with BaalChIP [69] and was called as an allele-specific binding site (ASB); (ii) the SNP and its surrounding sequence mapped into a biologically-relevant binding motif for the investigated TF; and, (iii) the direction of change in binding inferred at this site was concordant with the direction in which it disrupted/strengthened the TF binding motif. All high-quality ASBs predicted by [23] were downloaded and filtered for only NHRs [81]. Overall, 9/86 of the transcription factors with high-quality ASBs were NHRs: VDR, ESR1, NR3C1, AR, HNF4A, HNF4G, NR2F1, NR2F2, and PGR. We further required that the alleles tested in each cell line by *baal-nf* matched the alleles tested in the UK Biobank. Numbers of high-quality ASBs per TF are shown in **Table 1**. All bQTLs investigated in this study can be found in **Supplementary Table S2**. Per-allele read counts for archaic hominins were acquired from https://bioinf.eva.mpg.de/jbrowse/. This information for bQTLs discussed here is shown in **Supplementary Table S6**.

#### Defining input facilitator sets for each NHR

Facilitator QTLs were defined by three potential mechanisms: (i) change in the NHR’s mRNA transcript abundance, represented by eQTLs; (ii) change in abundance of the NHR-relevant hormone (hQTLs); and, (iii) change in SHBG protein abundance for mechanisms including two sex hormones, testosterone and estradiol (globQTLs).

Facilitatory QTLs were either (1) curated from the literature or (2) acquired from publicly-available sources GWAS Catalogue, eQTLGen and GTEx. GWAS catalogue was used to determine sets of hormone QTLs and globulin QTLs (protein QTLs for SHBG). For eQTLs, we used the set discovered in Whole Blood from eQTLGen if available, and if not then we deferred to eQTLs from GTEx (across all tissues). All final sets were clumped so that the final set of transactors per-TF contained no two SNPs in high LD with one another (*R*^2^ *<* 0.8). Details of final transactor sets across all 9 NHRs is provided in **Supplemental Table S1**.

### Pruning facilitator QTL sets for high Linkage Disequilibrium

For each set of facilitator QTLs, we derived a set whose SNPs are not in high LD. We first called genotypes across our population in the UK Biobank for all fQTLs and bQTLs from genotyped and imputed SNPs, requiring SNPs to be (i) biallelic, (ii) have an imputation quality score *>* 0.9 and (iii) minor allele frequency (MAF) *>* 0.01. SNP statistics were generated, and genotypes called using QCTool v2.0.8. The resulting BED files were merged using PLINK v1.90b7.2 and fQTLs further clumped with PLINK using flags `--clump-p1 5e-6 --clump-p2 0.05 --clump-r2 0.8`. This defined clumps of SNPs correlated at *R*^2^ *>* 0.8. In each clump, the most significant SNP was chosen to represent that haplotype block, where significance was the p-value calculated by the original eQTL, hQTL or globQTL study.

#### Defining LD blocks

Given our final set of bQTLs and fQTLs, we defined genomic regions whose variants are highly correlated with our SNPs-of-interest (*i*.*e*., LD blocks). These were computed across all genotyped SNPs for our population-of-interest in the UK Biobank, using QCtool v2.0.8. We scanned the region ± 10, 000, 000 bases around each investigated SNP (either bQTL or fQTL) and defined LD blocks by requiring a genomic region to have *R*^2^ *>* 0.05 with our SNP-of-interest. To do so, we used the following flags in QCtool v2.0.8: `-compute-ld-with and -min-r2 0.05`. This procedure provided LD blocks defined by upper and lower bounds.

#### Genetic confounders per TF

Given LD blocks, we computed confounders (*W* ), *i*.*e*., population stratification in the cohort. To do so, we removed regions defined by our LD blocks across all bQTLs and fQTLs from the genotyped SNPs in our population using the filter-chromosome functionality from TargeneCore v0.10.1. We further thinned by LD using PLINK v2.00a2.3LM and the flags `--indep-pairwise 1000 50 0.05` and also removed problematic regions of the genome, as recommended and provided by [82] using the flags `--exclude range exclusion_regions_hg19.txt`. We further filtered SNPs on Hardy-Weinberg equilibrium and missingness using PLINK2 and flags `--hwe 1e-10 --geno --mind`. On all remaining filtered, pruned, and quality controlled SNPs, we ran FlashPCA2 for up to 50 dimensions across all individuals in our cohort, and then visualized the number of PCs sufficient to capture population stratification using a ScreePlot. This procedure was performed for each TF (Fig. S1).

### Epistasis estimation with TarGene

Our aim in this work was to detect epistatic variant-variant interactions with maximal power and minimal bias. We thus required an estimation approach that is independent of parametric assumptions, unlike LMMs, that maximises power, such as DeepNull/XGBoost, yet benefits from mathematical guarantees of desirable statistical properties, such as minimal bias, consistency and 95% coverage of ground truth. We therefore imported and adapted semi-parametric estimation methods into population genetics and created a methodology and software, Targeted Genomic Estimation (TarGene) [27, 28], for the estimation of genetic effects. TarGene is based on techniques from the semi-parametric estimation literature, including Targeted Learning [25] and One-Step Estimation, which have been extensively developed and successfully applied in the last decade to problems in epidemiology, econometrics, health informatics, and public health policy [12, 26, 83].

We summarise the core methodological components of TarGene. TarGene leverages estimators that are asymptotically unbiased, normally distributed, and have minimum variance. Effect size estimation within this framework proceeds in two stages and involves several key modelling choices [25, 83]. These were guided by empirical performance across diverse simulation scenarios relevant to population genetics, including models with interaction effects [27, 28].

In the first stage, we estimate two nuisance functions using machine learning: the outcome regression and the propensity score [84]. For example, when modelling a potential interaction between a bQTL and an fQTL, adjusting for population stratification and additional covariates such as sex and age, these functions correspond to 𝔼[*Y* |bQTL, fQTL, PCs, Sex, Age] and *P* (bQTL, fQTL|PCs). We use XGBoost [85] to estimate these components, with cross-validation over tree depth and *L*_2_ regularisation strength to mitigate overfitting.

In the second stage, we apply Targeted Maximum Likelihood Estimation (TMLE) to refine initial estimates using appropriate clever covariates and weights, thereby reducing bias in the final effect size estimates [25]. Each targeting step yields an estimate for a specific genotype change (*e*.*g*., bb → bB or ff → fF). However, rather than testing individual genotype transitions, it is often more biologically meaningful to ask whether a given pair of loci, such as a bQTL and fQTL, interact. Because the TMLE estimators are asymptotically normal, these genotype-level estimates can be jointly analysed at the variant level using a Hotelling’s *T* ^2^ test [27]. Accounting for the correlation structure among genotype changes improves statistical power compared to testing each transition independently (see Fig. S4 and Section A.2 in Supplementary Material). Finally, as is common in GWAS, we impose a minimum frequency constraint to ensure robustness. Specifically, we exclude any genotype change with a minor genotype frequency below 0.01. This threshold ensures that each bQTL–fQTL interaction comprises at most four interpretable components.

## Supporting information

Main Tables

Table S1: All transactors

Table S2: All bQTLs

Table S3: All 2-point allelic effects

Table S4: PGR AR GxGxG results

Table S5: All 3-point allelic effects

Table S6: SNP evolution

Table 1: summary of investigated QTLs

Table 2: 2-point replicated interactions

Table 3: 3-point replicated interactions

## Supplementary information

All supplementary tables can be downloaded from the Open Science Framework at https://osf.io/wcx8e/ under the project osf.io/wcx8e.

## Declarations

### Funding

**BRH** was supported by the Health Data Research UK & The Alan Turing Institute Wellcome PhD Programme in Health Data Science (Grant Ref: 218529/Z/19/Z). **OL** was supported by the United Kingdom Research and Innovation (grant EP/S02431X/1), UKRI Centre for Doctoral Training in Biomedical AI at the University of Edinburgh, School of Informatics. **MvdL** is supported by NIH grant R01AI074345-09. **CPP** was supported by the Medical Research Council (grant MC UU 00007/15). **AK** was supported by the XDF Programme from the University of Edinburgh and Medical Research Council (MC UU 00009/2), and by a Langmuir Talent Development Fellowship from the Institute of Genetics and Cancer, and a philanthropic donation from Hugh and Josseline Langmuir.

### Conflict of interest/Competing interests

The authors declare no conflicts of interest or competing interests.

### Ethics approval and consent to participate

This research has been conducted using the UK Biobank Resource under Application Number 91924. UK Biobank has approval from the North West Multi-centre Research Ethics Committee (MREC) as a Research Tissue Bank (RTB) approval (2011, renewed 2016 and 2021). This approval means that researchers do not require separate ethical clearance and can operate under the RTB approval. The basis for consent in UK Biobank rests on participants’ explicit and informed consent. UK Biobank uses “legitimate interests” as the primary lawful basis on which to process personal data under the UK GDPR.

### Consent for publication

Not applicable.

### Data availability

This research was conducted using the UK Biobank Resource under Application Number 91924, and all data was made available to our group under this approved project. All relevant data to the research outputs described in this manuscript are contained in the main and supplementary tables. The input bQTLs and fQTLs were publicly available, provided by the manuscript [23] and publicly available sources GTEx [40], GWAS Catalog [86] and eQTLGen[37]. For the purpose of open access, the author has applied a creative commons attribution (CC BY) license to any author accepted manuscript version arising.

As stated above, all results generated in this manuscript can be downloaded from the Open Science Framework (OSF) at https://osf.io/wcx8e/ under the project osf.io/wcx8e. This includes all main Tables, Supplementary Tables as well as individual allelic effect plots for every significant interaction discovered in this study.

### Code availability

All code is publicly available on GitHub at the following repositories: (1) [TarGene Pipeline](https://github.com/TARGENE/targene-pipeline) which was used to estimate genetic interaction effects using the Targeted learning framework, (2) LD-block-removal Pipeline for computing LD blocks around our SNPs-of-interest, (3) [LD-clumping Pipeline](https://github.com/roskamsh/ld-clump/) for pruning fQTLs and determining the number of PCs required to estimate *W* . All code and run configurations for analyses carried out in this study can be found at [NHR-repository](https://github.com/edbiomedai/nhr).

### Author contributions

**BRH**: Formal analysis; Software; Investigation; Data curation; Writing - Original Draft; Writing - Review and editing; Visualization. **OL**: Methodology; Software; Formal analysis; Investigation; Writing - Review and Editing. **KTC**: Data curation; Software; Investigation. **MvdL**: Methodology, Writing - Review and editing. **CPP**: Conceptualisation; Investigation; Methodology; Supervision; Writing — Original draft; Project administration; Writing - Review and editing. **SB**: Conceptualisation; Investigation; Methodology; Formal Analysis; Software; Supervision; Writing - Original draft; Writing - Original draft; Project administration; Writing - Review and editing. **AK**: Conceptualisation; Investigation; Methodology; Formal Analysis; Software; Supervision; Writing - Original draft; Project administration; Writing - Review and editing.

## Acknowledgments

We would like to thank all participants of the UK BioBank without whom this study would not have been possible. Furthermore, we would like to thank Øyvind Almelid, John Ireland and Angela Chitzanidi for their input and help running these analyses on the University of Edinburgh high performance compute cluster, Eddie.

## Notes

### Competing Interest Statement

The authors have declared no competing interest.

