## Supplementary Material for "Epistatic contributions to human traits via transcription factor mechanisms"

[1*]MRC Human Genetics Unit, Institute of Genetics and Cancer, University of Edinburgh, Edinburgh, EH4 2XU, United Kingdom.
[2]Health Data Research UK, 215 Euston Road, London, NW1 2BE, United Kingdom.
[3]Institute for Regeneration and Repair, University of Edinburgh, 4–5 Little France Drive, Edinburgh, EH16 4UU, United Kingdom.
[4]School of Public Health, University of California, Berkeley, 2121 Berkeley Way, California, 94720, United States of America.
[5]School of Mathematics and Maxwell Institute for Mathematical Sciences, University of Edinburgh, Peter Guthrie Tait Road, Edinburgh, EH9 3FD, United Kingdom.
[6]School of Informatics, University of Edinburgh, 10 Crichton Street, Edinburgh, EH8 9AB, United Kingdom.

Contributing authors:;
[†]These authors contributed equally to this work.
[‡]These authors also contributed equally to this work.

**Keywords:** Epistasis, population genetics, functional genomics, nuclear hormone receptors, higher-order interactions, semi-parametric estimation

### A Tables

| Nuclear Hormone Receptor | Gene Symbol | # bQTLs | # fQTLs | Hormone | Protein |
|---|---|---|---|---|---|
| Androgen receptor | AR | 8 | 14 hQTLs; 21 globQTLs | Testosterone | Sex Hormone Binding Globulin |
| Progesterone receptor | PGR | 1 | 9 hQTLs; 32 eQTLs | Progesterone | |
| Estrogen receptor | ESR1 | 77 | 6 hQTLs; 21 globQTLs; 4 eQTLs | Estradiol | Sex Hormone Binding Globulin |
| Glucocorticoid receptor | NR3C1 | 10 | 2 hQTLs; 11 eQTLs | Cortisol | |
| Vitamin D receptor | VDR | 12 | 8 hQTLs; 2 eQTLs | 25-hydroxyvitamin D | |
| Nuclear receptor subfamily 1 | NR2F1 | 2 | 16 eQTLs | | |
| Nuclear receptor subfamily 2 | NR2F2 | 1 | 3 eQTLs | | |
| Hepatocyte nuclear factor 4$\alpha$ | HNF4A | 3 | 5 eQTLs | | |
| Hepatocyte nuclear factor 4$\gamma$ | HNF4G | 5 | 7 eQTLs | | |

**Table 1**: **Summary of the QTLs studied for this manuscript.** For each of 9 NHRs, we report the number of high-quality bQTLs, as well as fQTLs whose mechanism is either by altered mRNA expression of the TF (eQTLs), altered ligand abundance directly by the hormone (hQTLs), or by Sex hormone binding globulin (globQTLs). Additional information on each SNP can be found in **Supplemental Table S1 and S2**.

**Table 2**: All replicated 2-point interactions across 6 NHRs

| bQTL | Outcome | Transcription factor | # Trans-actors |
|---|---|---|---|
| rs2553234 | M86-M90 Other osteopathies | AR | 2 |
| rs8180759 | N93 Other abnormal uterine and vaginal bleeding | AR | 2 |
| rs4977574 | Poultry intake | AR | 2 |
| rs12626817 | I26-I28 Pulmonary heart disease and diseases of pulmonary circulation | PGR | 2 |
| rs12626817 | Part of a multiple birth | PGR | 2 |
| rs12626817 | angina | PGR | 2 |
| rs12626817 | M70-M79 Other soft tissue disorders | PGR | 2 |

Continued on next page

**Table 2 – continued from previous page**

| bQTL | Outcome | Transcription Factor | # Transactors |
|---|---|---|---|
| rs12626817 | B95 Streptococcus and staphylococcus as the cause of diseases classified to other chapters | PGR | 2 |
| rs12626817 | osteopenia | PGR | 2 |
| rs12626817 | J92 Pleural plaque | PGR | 2 |
| rs12626817 | cervical problem | PGR | 3 |
| rs12626817 | J34 Other disorders of nose and nasal sinuses | PGR | 3 |
| rs12626817 | gall bladder disease | PGR | 9 |
| rs12447081 | transient ischaemic attack (tia) | ESR1 | 2 |
| rs17708638 | Arm fat mass (left) | ESR1 | 3 |
| rs17708638 | Arm fat mass (right) | ESR1 | 3 |
| rs6470263 | O80 Single spontaneous delivery | HNF4A | 2 |
| rs6470263 | hayfever/allergic rhinitis | HNF4A | 2 |
| rs514795 | H25-H28 Disorders of lens | HNF4A | 2 |
| rs514795 | H25 Senile cataract | HNF4A | 2 |
| rs514795 | eye trauma | HNF4A | 2 |
| rs6470263 | uterine problem | HNF4A | 2 |
| rs1191818 | haematology | HNF4A | 2 |
| rs6470263 | J35 Chronic diseases of tonsils and adenoids | HNF4A | 3 |
| rs1191818 | anaemia | HNF4A | 3 |
| rs568557 | clotting disorder/excessive bleeding | HNF4G | 2 |
| rs514795 | I89 Other non-infective disorders of lymphatic vessels and lymph nodes | HNF4G | 2 |
| rs568557 | osteopenia | HNF4G | 2 |
| rs568557 | heart arrhythmia | HNF4G | 2 |
| rs514795 | G56 Mononeuropathies of upper limb | HNF4G | 3 |
| rs568557 | M07 Psoriatic and enteropathic arthropathies | HNF4G | 3 |
| rs6470263 | L57 Skin changes due to chronic exposure to nonionising radiation | NR2F1 | 2 |
| rs11642445 | K55-K64 Other diseases of intestines | NR2F1 | 2 |
| rs11642445 | neck problem/injury | NR2F1 | 2 |
| rs6470263 | Lymphocyte percentage | NR2F1 | 2 |
| rs11642445 | K04 Diseases of pulp and periapical tissues | NR2F1 | 2 |
| rs6470263 | viral infection | NR2F1 | 2 |
| rs11642445 | Hot drink temperature | NR2F1 | 2 |
| rs6470263 | E20-E35 Disorders of other endocrine glands | NR2F1 | 3 |
| rs6470263 | N32 Other disorders of bladder | NR2F1 | 3 |
| rs6470263 | L55-L59 Radiation-related disorders of the skin and subcutaneous tissue | NR2F1 | 3 |
| rs6470263 | Coffee intake | NR2F1 | 3 |
| rs11642445 | M77 Other enthesopathies | NR2F1 | 3 |
| rs6470263 | chronic obstructive airways disease/copd | NR2F1 | 4 |
| rs6470263 | J40-J47 Chronic lower respiratory diseases | NR2F1 | 4 |
| rs6470263 | F00-F09 Organic, including symptomatic mental disorders | NR2F1 | 4 |
| rs6470263 | M77 Other enthesopathies | NR2F1 | 4 |
| rs6470263 | F05 Delirium, not induced by alcohol and other psychoactive substances | NR2F1 | 5 |
| rs11642445 | Immature reticulocyte fraction | NR2F1 | 5 |
| rs6470263 | K42 Umbilical hernia | NR2F1 | 6 |

Continued on next page

Table 2 – continued from previous page

| bQTL | Outcome | Transcription Factor | # Trans-actors |
|---|---|---|---|
| rs11642445 | K31 Other diseases of stomach and duodenum | NR2F1 | 7 |
| rs11642445 | muscle or soft tissue injuries | NR2F1 | 8 |
| rs998384 | High light scatter reticulocyte count | NR2F2 | 2 |
| rs998384 | Leg fat percentage (left) | NR2F2 | 2 |
| rs998384 | L30 Other dermatitis | NR2F2 | 2 |
| rs998384 | Mean platelet (thrombocyte) volume | NR2F2 | 2 |
| rs998384 | G40 Epilepsy | NR2F2 | 2 |
| rs998384 | Beef intake | NR2F2 | 3 |

**Table 3**: All replicated 3-point interactions across 2 NHRs

| bQTL | Outcome | Transcription factor | # Trans-actors |
|---|---|---|---|
| rs4977574 | Variation in diet | AR | 2 |
| rs12626817 | K56 Paralytic ileus and intestinal obstruction without hernia | PGR | 2 |
| rs12626817 | G47 Sleep disorders | PGR | 2 |
| rs12626817 | B97 Viral agents as the cause of diseases classified to other chapters | PGR | 2 |
| rs12626817 | Monocyte count | PGR | 3 |
| rs12626817 | C15-C26 Malignant neoplasms of digestive organs | PGR | 3 |
| rs12626817 | L30 Other dermatitis | PGR | 3 |
| rs12626817 | L20-L30 Dermatitis and eczema | PGR | 3 |
| rs2286519 | Mean reticulocyte volume | ESR1 | 2 |
| rs1881561 | I60-I69 Cerebrovascular diseases | ESR1 | 3 |

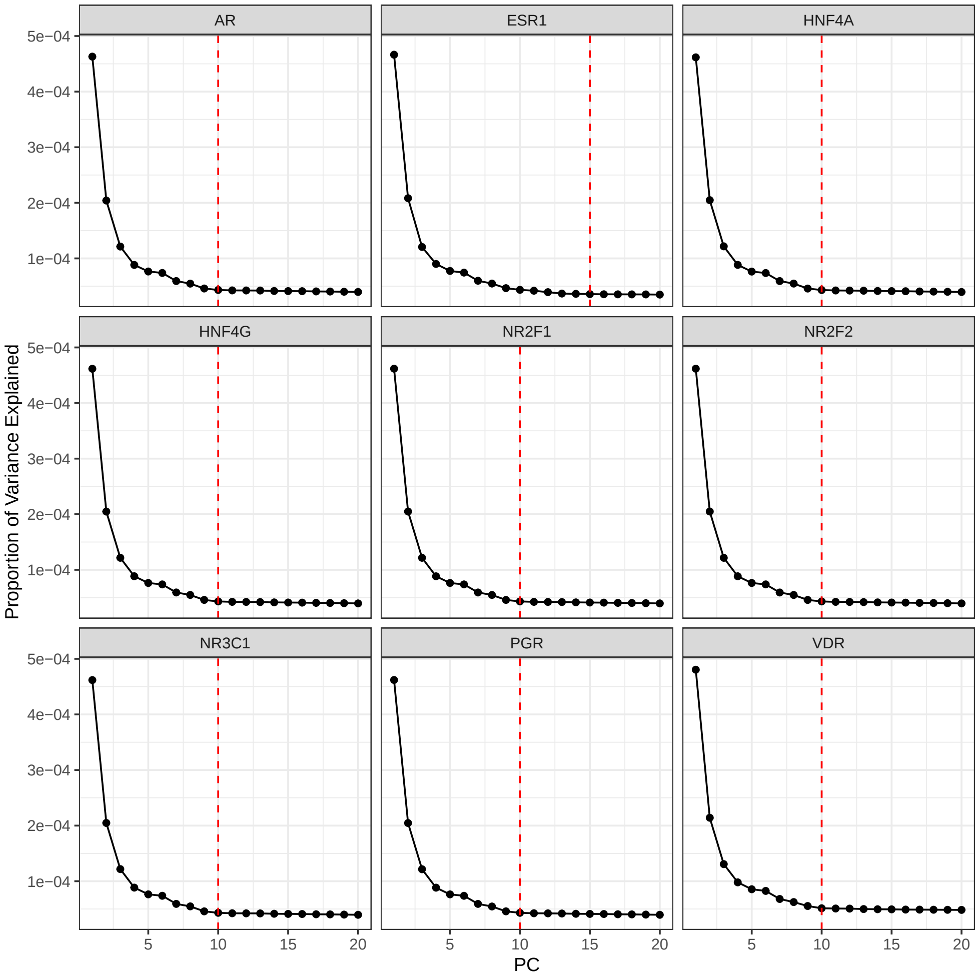


**Fig. S1**: **Scree plots across all 9 NHRs**. X-axes are the principal components up to 20, and the y-axis is the proportion of variance explained by these PCs. These plots were used to define the NHR-specific threshold for our confounders, **W**. Red dashed line indicates the cutoff used for each TF.

### B Supplemental figures

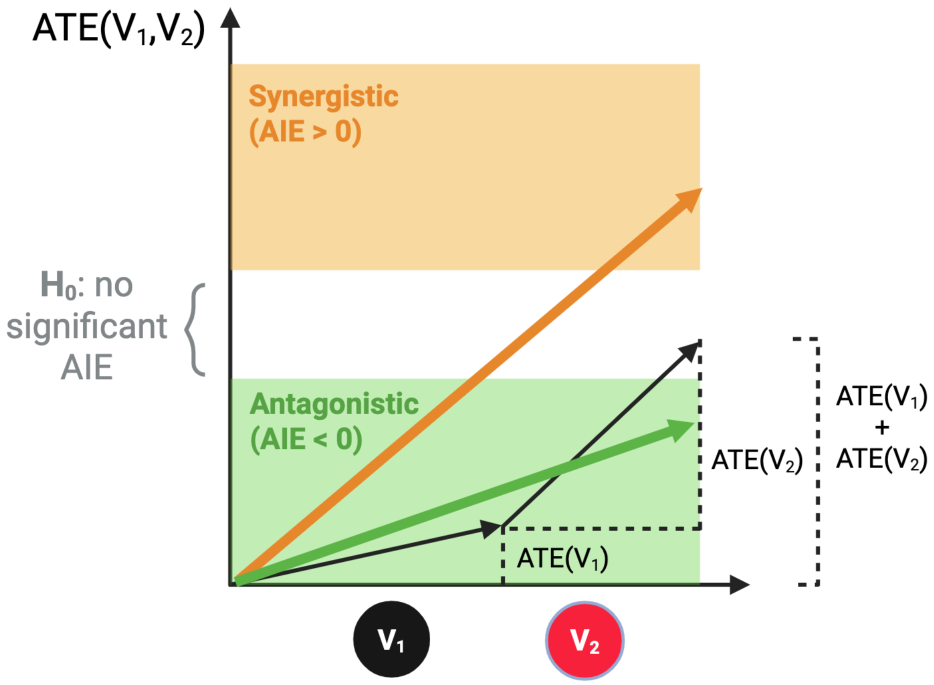


**Fig. S2**: **Schematic demonstrating synergistic or antagonistic epistasis.** The x-axis denotes changes in $V_1$, the bQTL, and $V_2$, the fQTL, and illustrates individual variant effects on trait measured by ATE. The sum of the individual effects of $V_1$ and $V_2$ on trait is denoted by $\text{ATE}(V_1) + \text{ATE}(V_2)$. The y-axis represents the effect on trait of a joint change in $V_1$ and $V_2$, $\text{ATE}(V_1, V_2)$. If the joint change in effect is larger than the sum of individual effects, the interaction is positive ($\text{AIE} > 0$) and said to be "synergistic" (top half; yellow). If the joint change in effect is smaller than the sum of individual effects, the interaction is negative ($\text{AIE} < 0$) and said to be "antagonistic" (lower half; green). If the joint change is not significantly different from the sum of the individual effects, the interaction is statistically zero (middle band; white).

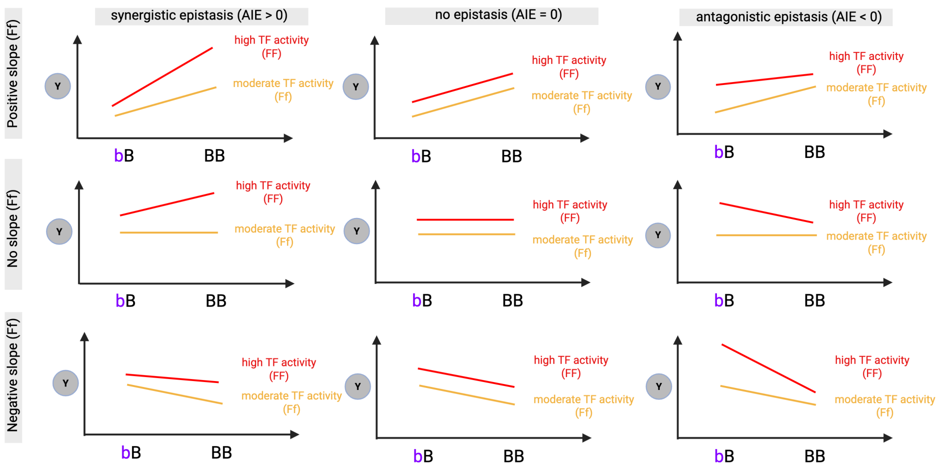


**Fig. S3**: **Various scenarios showing antagonistic and synergistic epistasis as a difference of conditional effects.** Varying scenarios displaying how antagonistic and synergistic epistasis could happen, when the gradients have the same sign (+/-). The three panels show scenarios where the effect of the bQTL at moderate fQTL (fF genotype) is positive (top row), zero (middle row) and negative (bottom row).

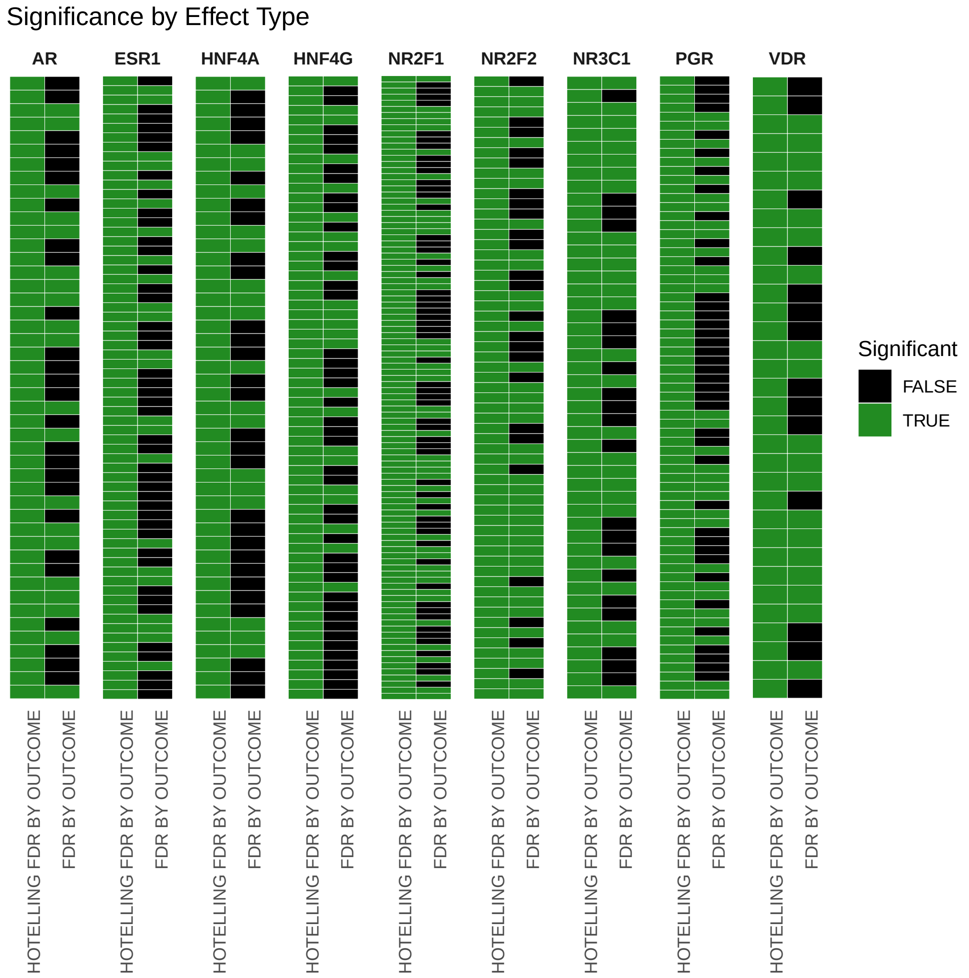


**Fig. S4**: **Advantage of Hotelling's $T^2$ statistic.** Here we show how many two-point interactions would be lost to correcting over all allelic changes, rather than correcting at the SNP-SNP-trait level. This demonstrates the advantage of using the Hotelling's $T^2$ test. Each block represents a NHR, with the left-hand column indicating whether the Hotelling FDR by outcome was significant, and the right-hand column representing whether the FDR across all allelic changes by outcome was significant. Each row represents a bQTL-fQTL-trait result for the two-point interaction runs. We note that across all NHRs, 292/535 (54.6%) interactions would not have been discovered had we corrected across all individual allelic changes.

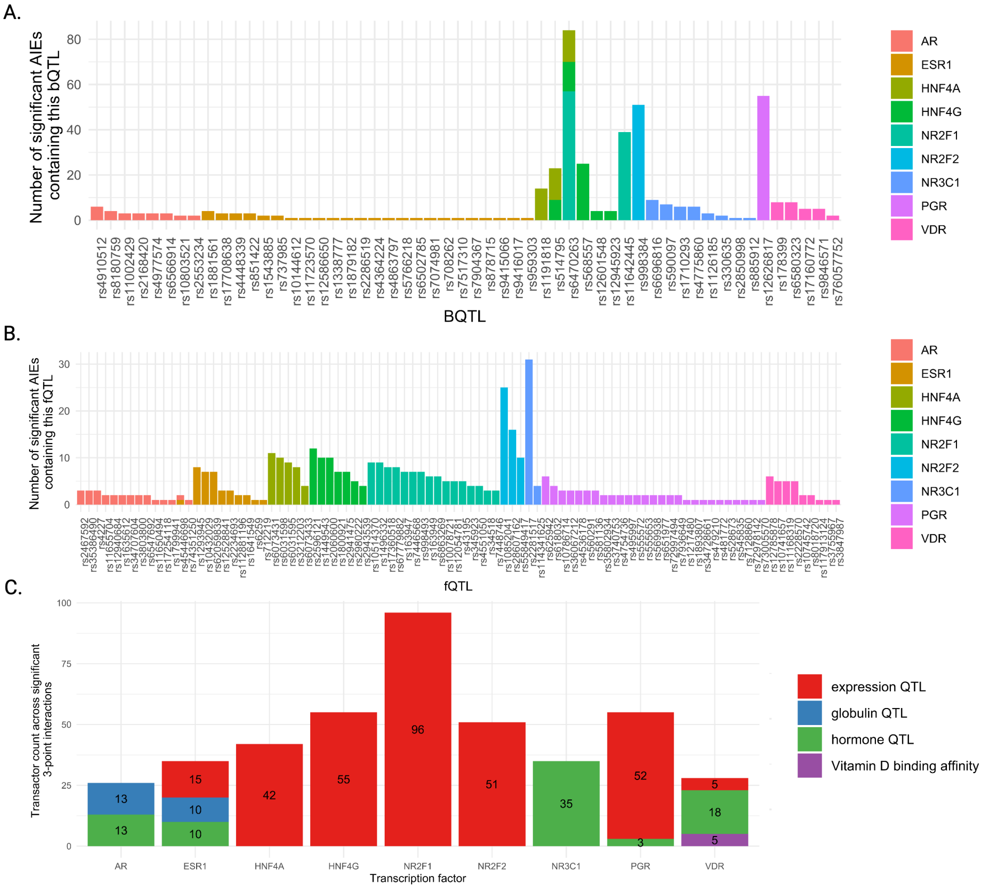


**Fig. S5**: **Summary of involved bQTLs, fQTLs and facilitator mechanisms in all 2-point interactions.** Counts for the number of **(A)** bQTLs, **(B)** fQTLs and **(C)** Transactor/fQTL mechanisms involved in all significant 2-point AIEs across 5 NHRs We note that there are 2 bQTLs shared among 3 transcription factors, HNF4A, HNF4G and NR2F1.

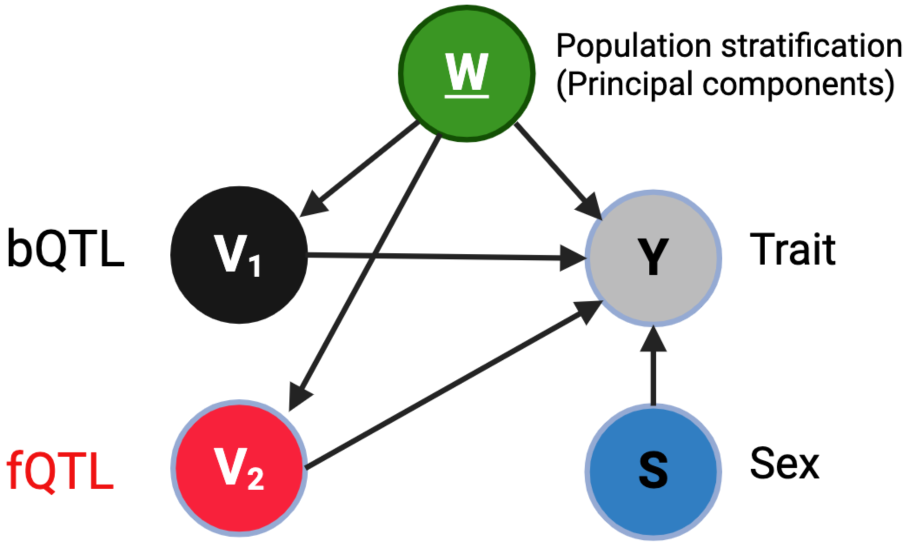


**Fig. S6**: **DAG for 3-point interactions including sex as an additional treatment.** As described for 2-point interactions in Fig. 1D, we also estimate 3-point interactions, where sex is used as an additional treatment. In this case, the treatment change would be represented by female → male alongside the respective genotype changes for the bQTL and fQTL.

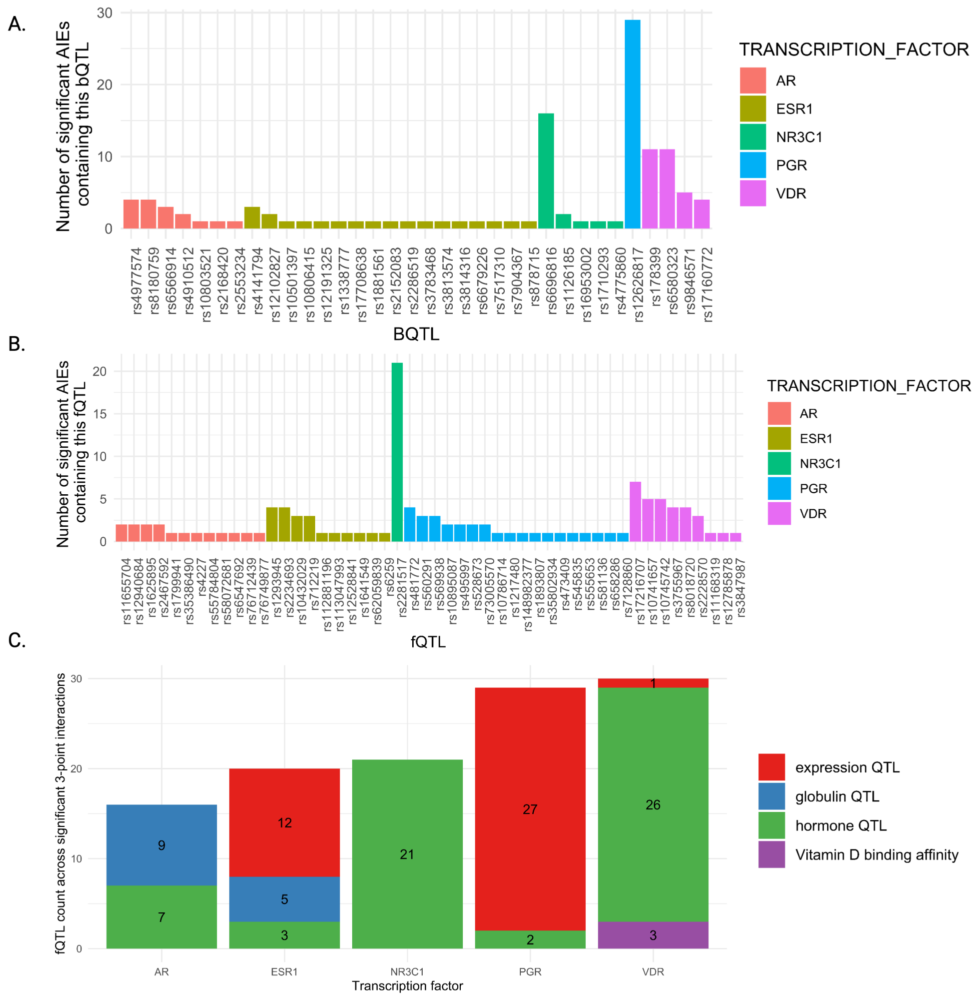


**Fig. S7**: **Summary of bQTLs, fQTLs and facilitator mechanisms involved in all 3-point interactions.** Counts for the number of **(A)** bQTLs, **(B)** fQTLs and **(C)** bQTL/fQTL mechanisms involved in all significant 3-point AIEs across 5 NHRs.

### C Supplemental methods

#### C.1 Statistical power calculation for ATE and AIE

In this section, we investigate the statistical power of detecting the marginal ATE of a variant $V_1$ on trait $Y$, compared to AIE between $V_1$ and $V_2$ on trait, at fixed total sample size $N$ (see Fig. 1A). We derive a closed-form expression for the statistical power in each case as a function of the sizes of these effects and genotype frequencies.

Under the null hypothesis of no effect, $H_0: \beta = 0$, the statistical power of the usual $Z$-test against the alternative hypothesis $H_1: \beta \neq 0$ of detecting a true effect size of $\beta$ (for ATE or AIE), is given by:

$$\text{Power} = P_{H_1}\left(Z > z_{1-\alpha} - \frac{\beta}{\sigma/\sqrt{N}}\right), \tag{1}$$

where $\sigma$ is the standard deviation of $Y$, $N$ is the sample size, $\sigma/\sqrt{N}$ is the standard error on the effect size estimate, $z_{1-\alpha}$ is the critical value for significance level $\alpha$, and $Z \sim \mathcal{N}(0,1)$ follows a standard normal distribution. For simplicity, in what follows, we will write $\beta$ in units of the standard error $\frac{\beta}{\sigma/\sqrt{N}} \to \beta$. At 95% confidence, *i.e.*, at $\alpha = 0.025$, the power can be written as $P(Z > 1.96 - \beta)$, and thus 80% power corresponds to $\beta = 2.8$ in units of its standard error.

For the purposes of illustration, in this example we assume that (i) $V_1$ and $V_2$ are independent, (ii) have binary genotype values with frequencies $f_1, 1-f_1$ and $f_2, 1-f_2$ respectively, (iii) there are no confounding variables involved, and (iv) the true causal model has the following form:

$$Y = \alpha_0 + \alpha_1 V_1 + \alpha_2 V_2 + \gamma V_1 V_2 + \epsilon, \tag{2}$$

where $Y$ is the trait, $\alpha_1$ and $\alpha_2$ are the pure biological effects of $V_1$ and $V_2$ on trait, $\gamma$ is the true biological interaction of $V_1$ and $V_2$ on trait, and $\epsilon \sim (0, \sigma^2)$ is a noise term. The sample average versions of Eq. 1 and Eq. 2,

$$\widehat{\text{ATE}}_{V_1:0\to1} = \frac{1}{N_1}\sum_{i:V_{1i}=1} Y_i - \frac{1}{N_0}\sum_{i:V_{1i}=0} Y_i \tag{3}$$

and

$$\widehat{\text{AIE}} = \frac{1}{N_{11}}\sum_{\substack{V_{1i}=1\\V_{2i}=1}} Y_i - \frac{1}{N_{10}}\sum_{\substack{V_{1i}=1\\V_{2i}=0}} Y_i - \frac{1}{N_{01}}\sum_{\substack{V_{1i}=0\\V_{2i}=1}} Y_i + \frac{1}{N_{00}}\sum_{\substack{V_{1i}=0\\V_{2i}=0}} Y_i, \tag{4}$$

are estimators of $\text{ATE}_{V_1:0\to1}$ and AIE ($\gamma$) with standard errors

$$\frac{\sigma}{\sqrt{N}}\frac{1}{\sqrt{f_1(1-f_1)}}, \quad \text{and} \quad \frac{\sigma}{\sqrt{N}}\frac{1}{\sqrt{f_1 f_2(1-f_1)(1-f_2)}}, \tag{5}$$

respectively. The relevant population sizes are $N_1 = Nf_1$ and $N_0 = N(1-f_1)$, whereas $N_{11} = Nf_1f_2$, $N_{10} = Nf_1(1-f_2)$, $N_{01} = N(1-f_1)f_2$, and $N_{00} = N(1-f_1)(1-f_2)$. In the special case where the ground truth biological ATE and AIE are equal in strength, and $f_1 = f_2 = 0.5$, the error on the ATE and AIE are $2\sigma/\sqrt{N}$ and $4\sigma/\sqrt{N}$ respectively. In other words, the error on the AIE is twice as large as ATE, and thus it would require 4 times the sample size to be able to detect AIE with the same statistical power as ATE, despite their true biological effect sizes being equal. This detection power will be lower for imbalanced genotype frequencies away from $f_2 = 0.5$, as is typically the case in population genetics, thus requiring an even larger sample size. These results are depicted in Fig. 1A (middle). Similarly, the relative power of detection is shown when the true magnitude of the AIE (blue) is half the ATE (left) and twice the ATE (right).

For AIE with a rare genotype frequency of $f_2 = 0.05$ to have the same detection power as ATE, one would require $\approx 21$ times the sample size, or its effect to be $\approx 4.59$ times as large as the ATE. Large-scale realistic simulation studies on UKB data performed in our earlier work (Labayle et al. 2025; Labayle 2025) confirm the expected drop in power when estimating AIE relative to ATE.

#### C.2 Significance testing using Hotelling's $T^2$ statistic

In this study, a two-point interaction of a bQTL-fQTL pair refers to a multidimensional effect arising from non-redundant combinations of allelic changes. For consistency and ease of interpretation, we considered the following 4 transitions,

$$\mathbf{AIE} = \begin{bmatrix} \text{AIE}_{(\text{bb},\text{ff})\to(\text{bB},\text{fF})} \\ \text{AIE}_{(\text{bb},\text{fF})\to(\text{bB},\text{FF})} \\ \text{AIE}_{(\text{bB},\text{ff})\to(\text{BB},\text{fF})} \\ \text{AIE}_{(\text{bB},\text{fF})\to(\text{BB},\text{FF})} \end{bmatrix} \tag{6}$$

which constitute all pairs of single-allelic changes corresponding to increased levels of binding and facilitator activity, respectively. In practice, only components for which genotypes are sufficiently frequent and can be estimated precisely (we require they pass the positivity criterion $\hat{p}(\text{bQTL}, \text{fQTL}) > 0.01$) are considered (Labayle et al. 2025; Labayle 2025). An estimate of our two-point interaction in Eq. 6 thus consists of $k \leq 4$ components, displayed in Fig. 1D. The proposed estimator for this quantity is asymptotically multivariate normal, allowing for a formal test of interaction between bQTL and fQTL using a Hotelling's $T^2$ test (Labayle et al. 2025; Labayle 2025). Using Hotelling's $T^2$ joint testing allows us to detect epistatic interactions at the DNA variant-level, rather than at the level of genotype changes. The resulting p-values (termed $p_H$) were adjusted using the Benjamini-Hochberg procedure and significant AIEs were defined at a false discovery rate (FDR) $< 0.05$ per trait per NHR (denoted in this study by $\text{FDR}_H$).

### D Recent methodological advances in population genetics

Statistical models in common use for association testing in population genetics include logistic, linear and linear mixed models (LMMs) (Uffelmann et al. 2021). LMMs are joint population-level models that incorporate variants, confounders and covariates as fixed effects, and population relatedness via the Genetic Relationship Matrix (GRM) as a random effect (Uffelmann et al. 2021; Svishcheva et al. 2012; Lippert et al. 2011). Another method, REGENIE (Mbatchou et al. 2021), takes a two-step ridge regression approach which avoids the need for the GRM by partitioning the genome into blocks, fitting an outcome regression on each block, and using the predictions as covariates in a subsequent association test through residual fitting. This reduces computational cost and memory burden.

These models, however, all rely on parametric assumptions, including linearity of the variant-covariate-trait relationship, and normality of the associated conditional distribution. Furthermore, since for any variant, trait, and confounder combination, the validity of any linearity assumption and optimality of power is a priori unknown, other methods such as DeepNull/XGBoost (McCaw et al. 2022) have been developed to data-adaptively account for genetic and confounder/covariate non-linearities. However, whilst these methods have been shown to increase power, they lack theoretical guarantees of consistency and 95% coverage of ground truth.

Most single variant genetic effect sizes are small and require large samples to be reliably estimated (Uffelmann et al. 2021). As demonstrated in Section C.1, the statistical detection power is even lower for AIE, even when their true effect size is of the same order of magnitude as the ATE. Therefore, a slight bias, such as when an estimation model is misspecified, can appreciably alter genetic effect estimates and, additionally, their associated confidence intervals may not have the theoretical nominal coverage of the ground truth. This problem of bias is exacerbated for larger sample sizes that yield more confident estimates (Labayle et al. 2025; Labayle 2025).

### E Causal versus statistical gaps

There are two main steps to causal inference: (i) Causal identification and (ii) statistical inference. Causal identification is the process of turning the causal quantity of interest (causal estimand) into a statistical quantity estimable from observed data (statistical estimand). Causal identification does not require data and is entirely separate from statistical inference. Statistical inference is the process of estimating the statistical quantity of interest, using statistical or machine learning models, together with uncertainty quantification. Causal identification proceeds by outlining assumptions about the causal relationships among treatments/interventions (here variants), confounders, covariates and outcomes of interest. These assumptions are typically encoded in a structural causal model (SCM) and graphically represented using causal diagrams known as directed acyclic graphs (DAGs) (Shrier and Platt 2008). The graph encodes the causal assumptions we are willing to make. Examining the structure of the DAG can identify sufficient sets of confounders to adjust for to derive a statistical estimand from the causal estimand (Pearl 2009; Hernan and Robins 2025). If a causal quantity is identifiable, algorithmic rules can be used to map the causal estimand into a statistical estimand. Note that given a graph, the causal effect of interest may not be identifiable, *i.e.*, it may not be estimable from observational data alone.

In the context of identifying variants that are causal of trait or disease, a suitable DAG, inspired by (Tudball et al. 2022), is presented in Figure S8. For the causal effect of genetic variants in Figure S8 to be identifiable, two main backdoor paths need to be blocked by adjusting: (i) variables corresponding to dynastic effects, whereby an individual's trait is directly influenced by their parent's phenotypes, and, (ii) genetic dependence across the genome inherited as dependent blocks during meiosis, and thus variants within the block are correlated (Berisa and Pickrell 2016). Unfortunately, in practice, neither can be fully blocked: (i) While family-based studies can mitigate bias from these confounding factors (Brumpton et al. 2020), most large-scale biobanks, such as UKB, lack extensive parental data, limiting our ability to adjust for these effects; and, (ii) Blocks of highly correlated variants can span up to several megabases, thereby obfuscating the causal variants from their linked counterparts. If variants are (almost) perfectly correlated, backdoor adjustment will lead to violation of the positivity assumption. Specifically, let $V_c = v_c$ be a candidate variant and genotype of interest, and let $\mathbf{V}_b$ be a set of linked variants in the block. For positivity to hold, it must be true that $p(V_c = v_c|\mathbf{V}_b = \mathbf{v}_b) > 0$, for all combinations $\mathbf{v}_b$. This constraint is almost impossible to achieve given the size of linkage disequilibrium blocks and the high degree of correlation within these blocks.

In general, the identification of causal variants through formal identification strategies remains a challenging and open research direction. While TarGene estimators address any statistical gap due to model misspecification as well as the causal gap due to population stratification, they do not currently close the causal gap due to LD. Attempts have been made in the statistics literature to address the causal gap due to LD through fine-mapping, *e.g.*, SuSiE (Wang et al. 2020; Zou et al. 2022), and KnockOffGWAS (Sesia et al. 2021). However, these methods do not close the statistical gap due to parametric assumptions or are unable to report (interaction) effect estimates, respectively. In this study, we have integrated information from functional genomics involving variants that are used as proxies for alteration of biological mechanisms, with population genetics, through the study of epistasis. TarGene is therefore

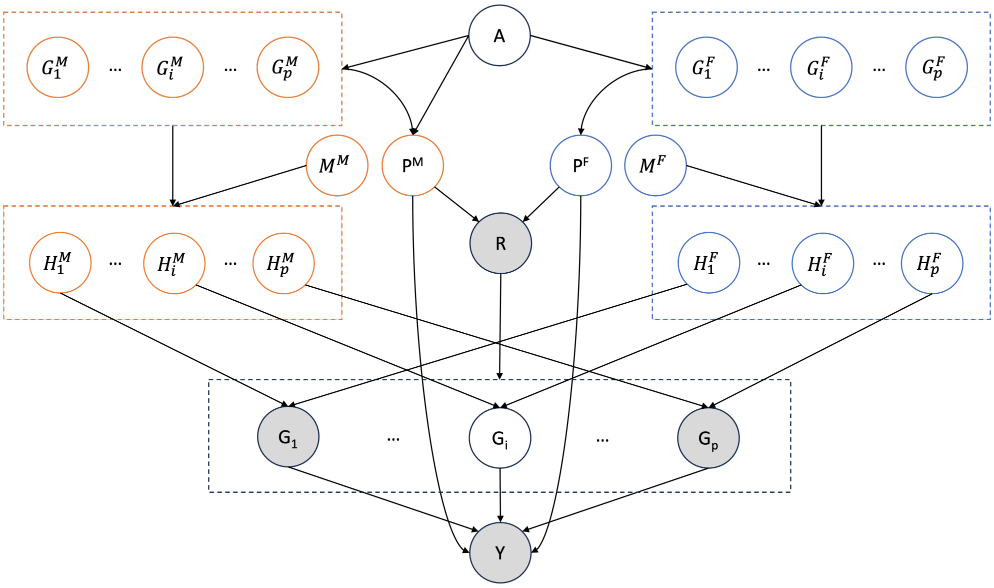


**Fig. S8**: **The proposed causal model of genetic inheritance.** Filled nodes represent observed variables in the UK Biobank while transparent nodes are unobserved. Unlike trio studies, in genotyped data not all genetic variations are observed, since the parental genotypes are unobserved. The displayed model captures how genetic variations and traits are inherited from one generation to the next. Linkage disequilibrium, dynastic effects and genetic ancestry confound genetic association studies.

the optimal methodology for this task, as it maximises power and minimises statistical bias via targeting variant interaction effects directly. This closes the statistical gap relative to the associational ground truth, as well as closes the causal gap due to population stratification. The causal gap due to LD is reduced in our approach, due to strong experimental evidence of altered bQTLs. A causal gap may however remain: (i) if there is a bQTL for another mechanism, in LD with the bQTL under examination, that also interacts with the transacting fQTL, or vice versa, or (ii) there is a bQTL in LD with another bQTL for the same mechanism, which is not prioritised as a 'high-quality' bQTL. If the former occurs, the incorrect mechanism may be prioritised. If the latter occurs, the mechanism would remain relevant, but the variant, or the cis-genes, may not be prioritised correctly depending on the strength of LD and/or proximity of the cis-genes.
